# The association between time-weighted remnant cholesterol and cardiovascular and non-cardiovascular mortality: A population-based cohort study

**DOI:** 10.1101/2024.02.08.24302507

**Authors:** Lifang Li, Vanessa Hou Cheng Chou, Oscar Hou In Chou, Sakshi Roy, Jeffrey Shi Kai Chan, Wing Tak Wong, Tong Liu, Gregory Lip, Bernard Man Yung Cheung, Gary Tse, Jiandong Zhou

## Abstract

**Background:** Remnant cholesterol (RC) have been suggested as a significant mediator of atherosclerotic cardiovascular diseases. However, the relationship between RC with cause-specific mortality in long-term remained uncertain. This study aimed to investigate the association between time-weighted RC and cause-specific mortality outcomes.

**Methods:** This retrospective population-based study enrolled patients attending family medicine clinics in Hong Kong between 1^st^ January 2000, to 31^st^ December 2003 with at least three RC testing results during follow-up. The time-weighted RC was calculated by the products of the sums of two consecutive measurements and the time interval divided by the total time. The primary outcomes were all-cause mortality and cause-specific mortality outcomes. Cox regression and marginal effective plots were applied to identify associations between time-weighted RC and mortality.

**Results:** A cohort of 75,342 patients (39.69% males, mean age: 61.3 years old) with at least three valid RC test were included. During up to 19 years of follow-up, in the multivariate model adjusted for demographics, comorbidities, medications, and time-weighted laboratory results, time-weighted RC was associated with all-cause mortality (Hazard ratio [HR]: 1.41; 95% Confidence Interval [CI]: 1.35-1.48) but not RC (HR: 0.99; 95% CI: 0.89-1.10). Time-weighted RC was also associated with increased risks of cardiovascular-related mortality (HR: 1.40; 95% CI: 1.27-1.54), cancer-related mortality (HR: 1.59; 95% CI: 1.43-1.77), and respiratory-related mortality (HR: 1.33; 95% CI: 1.20-1.47). The exploratory analysis of the cause of death demonstrated that time-weighted RC was associated with Ischaemic heart disease, cerebrovascular-related and pneumonia.

**Conclusions:** Time-weighted RC was independently associated with all-cause mortality and cause-specific mortality outcomes amongst the general population.

## Introduction

Cardiovascular disease (CVD) remains a formidable adversary in global health, accounting for a significant proportion of morbidity and mortality worldwide (1). Despite understanding in traditional lipid profiles, particularly low-density lipoprotein cholesterol (LDL-C), a residual risk of cardiovascular events persists. Remnant cholesterol (RC), the cholesterol content in triglyceride-rich lipoproteins (TGRLs), emerges as a pivotal factor in this context(2). The relationship between RC and cardiovascular outcomes has been a subject of extensive study, revealing its significant association with atherosclerosis and total mortality(3). However, the intricacies of this relationship, particularly in the context of time-weighted RC levels and their association with cause-specific mortality, remain less explored(4).

Furthermore, the association of RC with non-cardiovascular mortality adds another layer of complexity to its clinical significance. While studies have predominantly focused on cardiovascular outcomes, the potential link between RC and other causes of mortality, such as non-alcoholic fatty liver disease, warrants comprehensive investigation(5). This study aims to bridge these gaps by examining the association between time-weighted RC and both cardiovascular and non-cardiovascular mortality in a population-based cohort. It seeks to provide a more detailed understanding of RC’s role in mortality risk and to inform future strategies for cardiovascular risk assessment and management. This study aimed to examine the association between time-weighted remnant cholesterol (RC) and specific causes of mortality in a large cohort. This research focuses on time-weighted RC to provide a dynamic picture of RC’s long-term effects on all-cause mortality. Our findings aim to improve comprehension of lipid-related risk factors and inform better cardiovascular and general health management strategies.

## Methods

This study was approved by the Institutional Review Board of the University of Hong Kong/ Hospital Authority Hong Kong West Cluster (Reference No. UW 20-250) and complied with the Declaration of Helsinki.

### Study design and population

This is a retrospective cohort study with prospectively collected clinical data of patients attending family medicine clinics in Hong Kong between 1^st^ January 2000, to 31^st^ December 2003. The patients were followed up until 31^st^ December 2019. The patients were identified from the Clinical Data Analysis and Reporting System (CDARS), a territory-wide database by the Hospital Authority (HA) of Hong Kong that centralizes anonymized patient information from individual local hospitals, and this system has been used by multiple teams in Hong Kong. The records cover both public hospitals and their associated outpatient clinics as well as ambulatory and day-care facilities in Hong Kong. The coding was performed by the physicians who were not involved in the model development. Prior comorbidities at baseline were extracted. International Classification of Diseases Ninth Edition (ICD-9) codes were used to identify prior comorbidities provided in **Supplementary Table 1**.

### Covariates and primary outcomes

According to Nordestgaard and Varbo (6), RC was calculated by total cholesterol minus LDL-C minus HDL-cholesterol (HDL-C). Of which the LDL-C was calculated as total cholesterol minus HDL-C minus triglycerides divided by 5 according to the Friedewald equation (7). The standard deviation of the variables was also calculated **(Supplementary Table 2)**. The time-weighted lipid and glucose profiles were calculated by the products of the sums of two consecutive measurements and the time interval, then divided by the total time interval. The estimated glomerular filtration rate (eGFR) was calculated using the abbreviated modification of diet in renal disease (MDRD) formula (8).

Mortality data were obtained from the Hong Kong Death Registry, a population-based official government registry with the registered death records of all Hong Kong citizens. Mortality was recorded using the International Classification of Diseases Tenth Edition (ICD-10) coding. The ICD-10 of the mortality outcome were defined according to previous published literature (9).

### Statistical analysis

Descriptive statistics were used to summarise the baseline and clinical characteristics of patients. Continuous variables were presented as median (95% confidence interval [CI] or interquartile range [IQR]) and categorical variables were presented as count (%). The Mann-Whitney U test was used to compare continuous variables. The χ2 test with Yates’ correction was used for 2×2 contingency data. Cohorts were divided according to stratified RC and time-weighted RC into four quantiles. Univariable and multivariable Cox regression were used to explore the possible relationships between RC and the outcomes. Marginal effect plots were used to delineate the relationship between the RC and the outcomes. Hazard ratios (HRs) with corresponding 95% CIs and P values were reported. All significance tests were two-tailed and considered significant if P values were equal to or less than 0.05. No imputation was performed for missing data. Restricted cubic spline models with 3 knots were used to investigate the associations of RC and time-weighted RC with cause-specific mortality. Data analyses were performed using R-Studio software (Version: 1.1.456), STATA (Version: 16.1) and Python (Version: 3.6).

## Results

### Basic characteristics

The cohort included a total of 155,066 patients who attended family medicine clinic between 1^st^ January 2000 until 31^st^ December 2003. After excluding non-adult patients, patients without baseline RC, and patients without at least three valid RC test results during follow-up visits, 75,342 patients (39.69% males, mean age: 61.3 years old [Standard deviation: 13.1] remained in the cohort.

Among the included cohort, 23475 (31.15%) patients developed all-cause mortality during the follow-up period. In particular, 4349 (5.77%) developed cancer-related mortality, 4533 (6.01%) developed cardiovascular-related mortality, and 2029 (10.73%) developed respiratory-related mortality. 2627 (3.48%) patients died due to unknown causes. The characteristics of patients were stratified into four time-weighted RC quantiles, presented in **Table 1**, and the characteristics and outcomes of patients stratified into RC quartiles were also included **(Supplementary Table 3-4)**. The relationship between RC and HDL-C, LDL-C, total cholesterol, and triglyceride on morality were illustrated in **Supplementary Figure 1**.

**Table 1.**
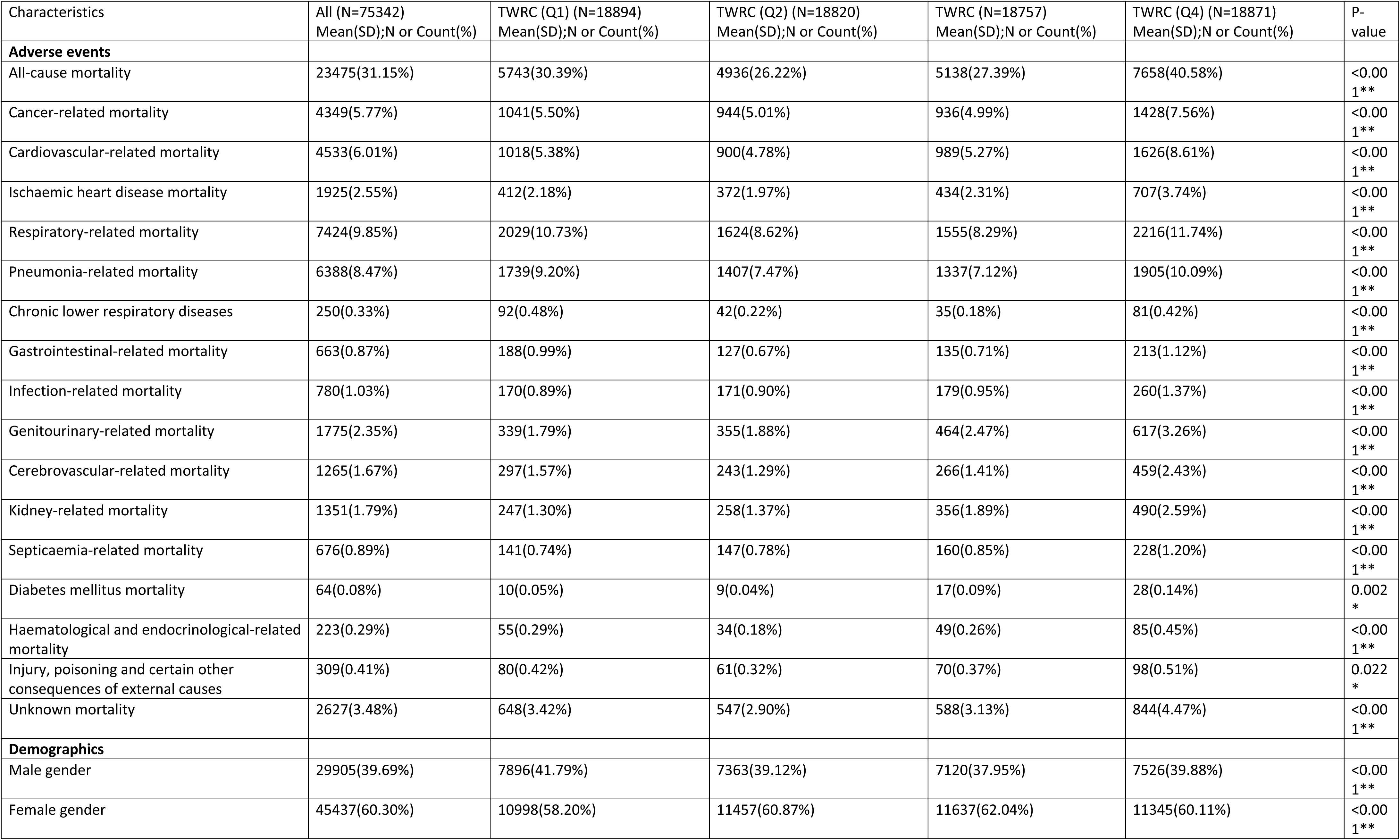
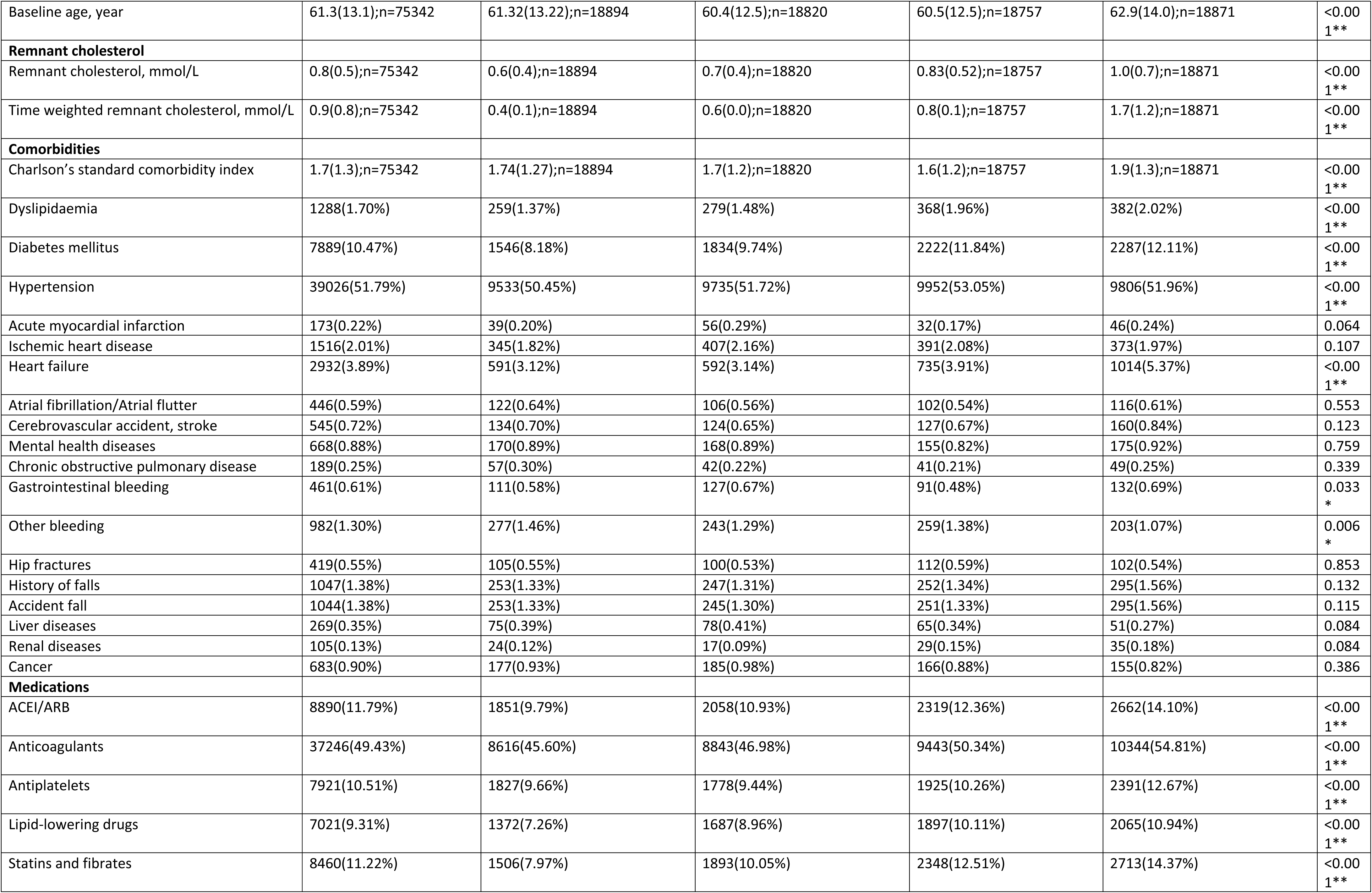
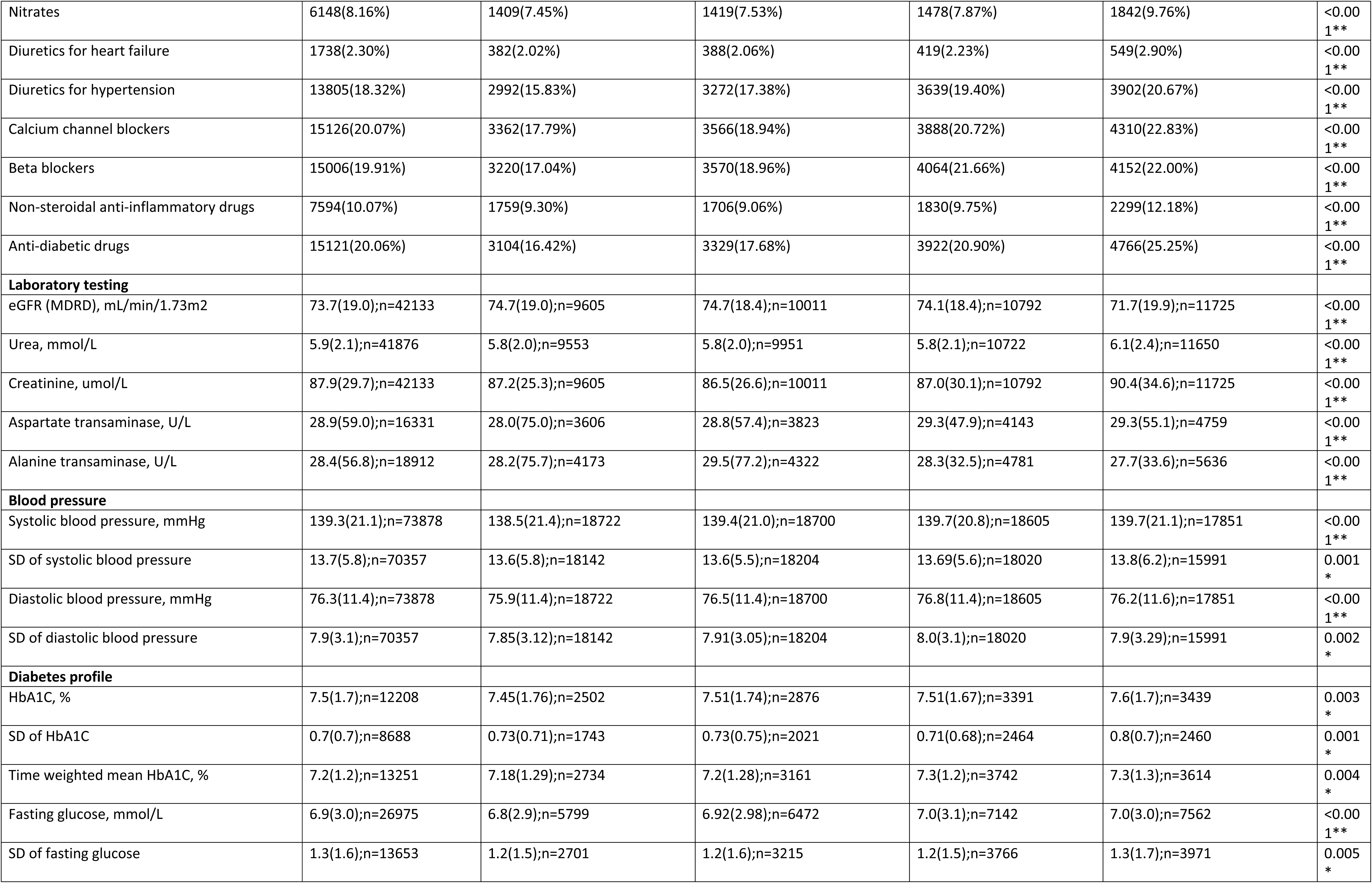
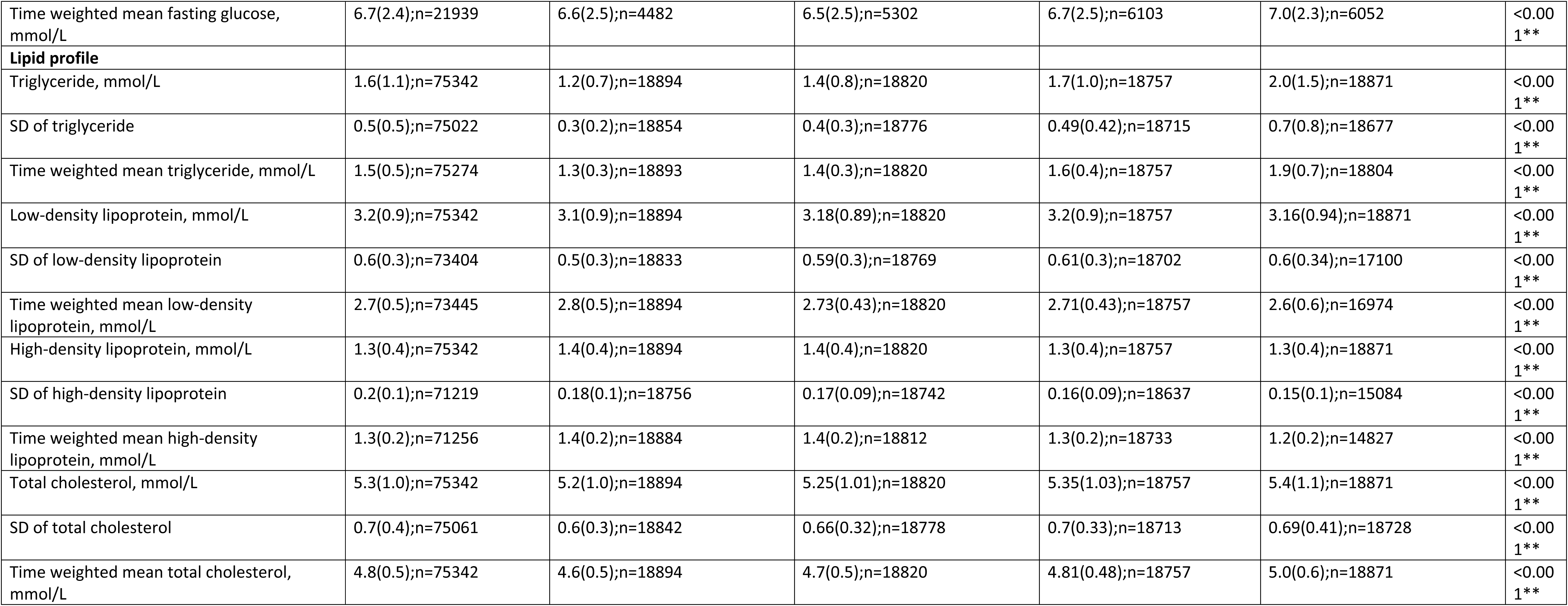
The number of adverse events, baseline, and clinical characteristics of patients in the study cohorts of four quartiles time-weighted remnant cholesterol. * for p≤ 0.05, ** for p ≤ 0.01, *** for p ≤ 0.001, TWRC: time-weighted remnant cholesterol, mmol/L, SD: standard deviation, ACEI: angiotensin-converting-enzyme inhibitors, ARB: angiotensin II receptor blockers, eGFR: estimated glomerular filtration rate.

**Table 2.** Multivariable Cox models assessing the relationships between remnant cholesterol and time-weighted cholesterol and cause-specific mortality. (Adjusted for demographics, comorbidities, medications, AMDRD, blood pressure, time-weighted lipid, and glucose tests). * for p≤ 0.05, ** for p ≤ 0.01, *** for p ≤ 0.001, HR: hazard ratio, CI: confidence interval.

| Characteristics | Number of patients (%) | Incidence per 1000 person-year [95%CI | Remnant cholesterol HR[95%CI];p-value | Time-weighted remnant cholesterol HR[95%CI];p-value |
| --- | --- | --- | --- | --- |
| All-cause mortality | 23475(31.15%) | 19.8[19.6- 20.1] | 0.99[0.89-1.10];0.8411 | 1.41[1.35-1.48];<0.0001*** |
| Cancer-related mortality | 4349(5.77%) | 3.7[3.6-3.8] | 0.86[0.64-1.14];0.2860 | 1.59[1.43-1.77];<0.0001*** |
| Cardiovascular-related mortality | 4533(6.01%) | 3.8[3.7-3.9] | 1.16[0.95-1.41];0.1475 | 1.40[1.27-1.54];<0.0001*** |
| Ischaemic heart disease mortality | 1925(2.55%) | 1.6[1.6-1.7] | 1.20[0.92-1.57];0.1817 | 1.28[1.10-1.48];0.0011** |
| Cerebrovascular-related mortality | 1265(1.67%) | 1.1[1.0-1.1] | 0.90[0.54-1.49];0.6728 | 1.47[1.20-1.79];0.0002*** |
| Respiratory-related mortality | 7424(9.85%) | 6.3[6.1-6.4] | 1.13[0.94-1.36];0.2075 | 1.33[1.20-1.47];<0.0001*** |
| Pneumonia-related mortality | 6388(8.47%) | 5.4[5.3-5.5] | 1.07[0.87-1.31];0.5366 | 1.33[1.20-1.48];<0.0001*** |
| Chronic lower respiratory diseases | 250(0.33%) | 0.2[0.2-0.2] | 2.31[1.13-4.75];0.224* | 1.68[1.12-2.54];0.0128* |
| Gastrointestinal-related mortality | 663(0.87%) | 0.6[0.5-0.6] | 0.64[0.31-1.35];0.2402 | 1.33[0.96-1.82];0.0829 |
| Infection-related mortality | 780(1.03%) | 0.7[0.6-0.7] | 0.89[0.52-1.53];0.6783 | 1.63[1.29-2.06];<0.0001*** |
| Genitourinary-related mortality | 1775(2.35%) | 1.5[1.4-1.6] | 0.99[0.74-1.31];0.9205 | 1.36[1.18-1.56];<0.0001*** |
| Kidney-related mortality | 1351(1.79%) | 1.1[1.1-1.2] | 1.10[0.82-1.47];0.5402 | 1.33[1.12-1.56];0.0008*** |
| Septicaemia-related mortality | 676(0.89%) | 0.6[0.5-0.6] | 0.95[0.55-1.64];0.8465 | 1.55[1.20-2.01];0.0009*** |
| Diabetes mellitus mortality | 64(0.08%) | 0.1[0.0-0.1] | 1.50[0.65-3.46];0.3380 | 1.37[0.83-2.27];0.2157 |
| Haematological and endocrinological-related mortality | 223(0.29%) | 0.2[0.2-0.2] | 0.89[0.45-1.77];0.7439 | 1.59[1.16-2.18];0.0036** |
| Injury, poisoning and certain other consequences of external causes | 309(0.41%) | 0.3[0.2-0.3] | 2.13[0.91-5.00];0.0831 | 2.13[1.47-3.07];0.0001*** |
| Unknown mortality | 2627(3.48%) | 2.2[2.1-2.3] | 0.73[0.51-1.04];0.0831 | 1.20[0.99-1.46];0.0678 |

### The association between remnant cholesterol and cause-specific mortality

The Multivariable Cox demonstrated that time-weighted RC (Hazard ratio [HR]: 1.41; 95% Confidence Interval [CI]: 1.35-1.48) but not RC (HR: 0.99; 95% CI: 0.89-1.10) was associated with increased risks of all-cause mortality after adjustments for demographics, comorbidities, medications, AMDRD, blood pressure, time-weighted lipid, and glucose tests. Amongst which, time-weighted RC was associated with increased risks of cardiovascular-related mortality (HR: 1.40; 95% CI: 1.27-1.54), cancer-related mortality (HR: 1.59; 95% CI: 1.43-1.77), and respiratory-related mortality (HR: 1.33; 95% CI: 1.20-1.47) following adjustments. The exploratory analysis of the cause of death subcategories demonstrated that time-weighted RC was associated with Ischaemic heart disease mortality (HR: 1.28; 95% CI: 1.10-1.48), cerebrovascular-related mortality (HR: 1.47; 95% CI: 1.20-1.79) and pneumonia-related mortality (HR: 1.33; 95% CI: 1.20-1.48). Time-weighted RC was also associated with other mortality outcomes apart from gastrointestinal-related mortality, diabetes mellitus mortality and unknown mortality. Meanwhile, RC was not associated with any mortality outcome (all p>0.05).

The cumulative curve also demonstrated that time-weighted RC had better discriminative power than RC for most of the mortality outcomes (**Figure 2**; **Supplementary Figure 2**). The marginal effect plots also demonstrated that as the time-weighted RC increased, the HR of the mortalities increases **(Figure 3; Supplementary Figure 3)**.

**Figure 1.**
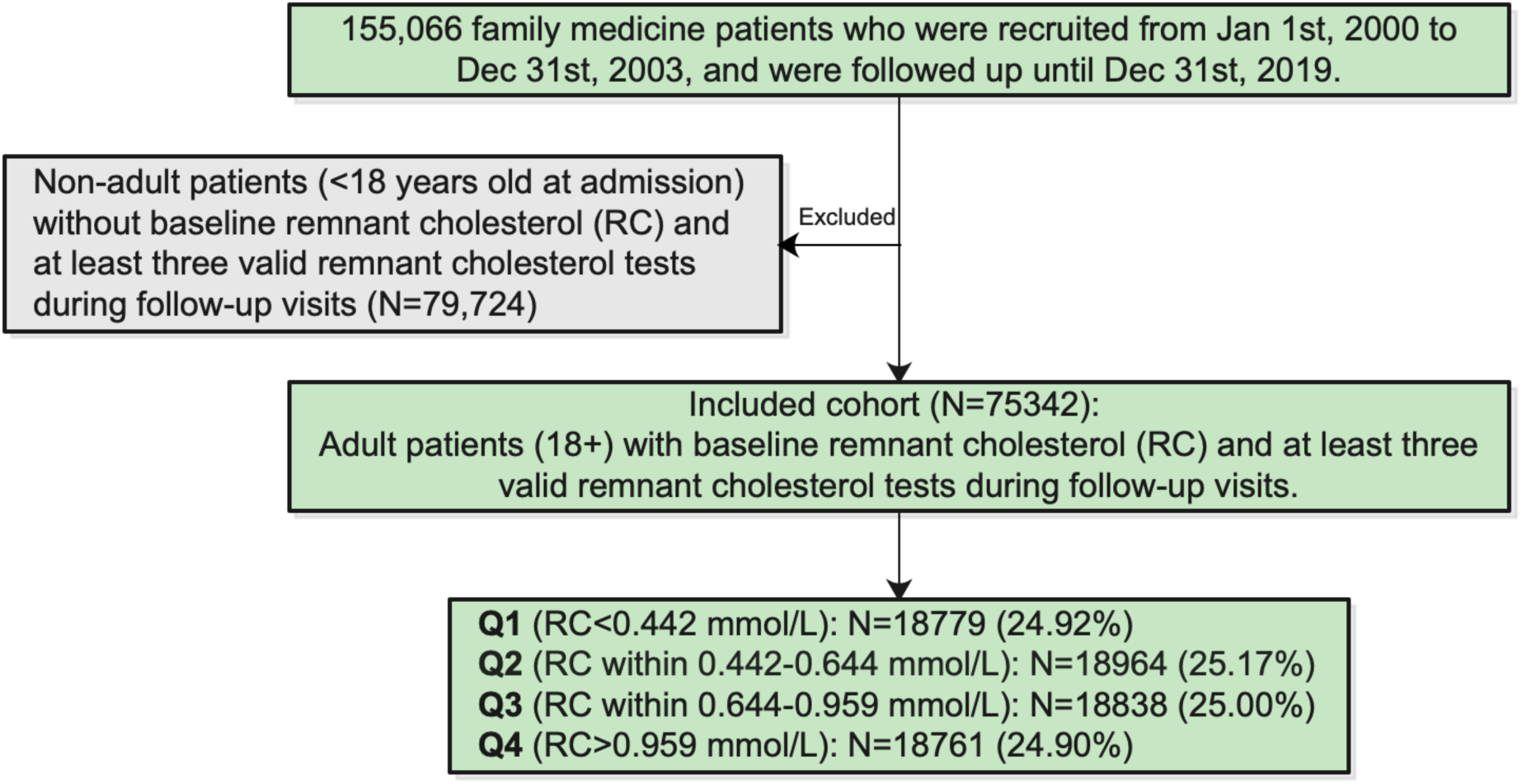
Flow chart for the identification, inclusion, and exclusion of study subjects.

**Figure 2.**
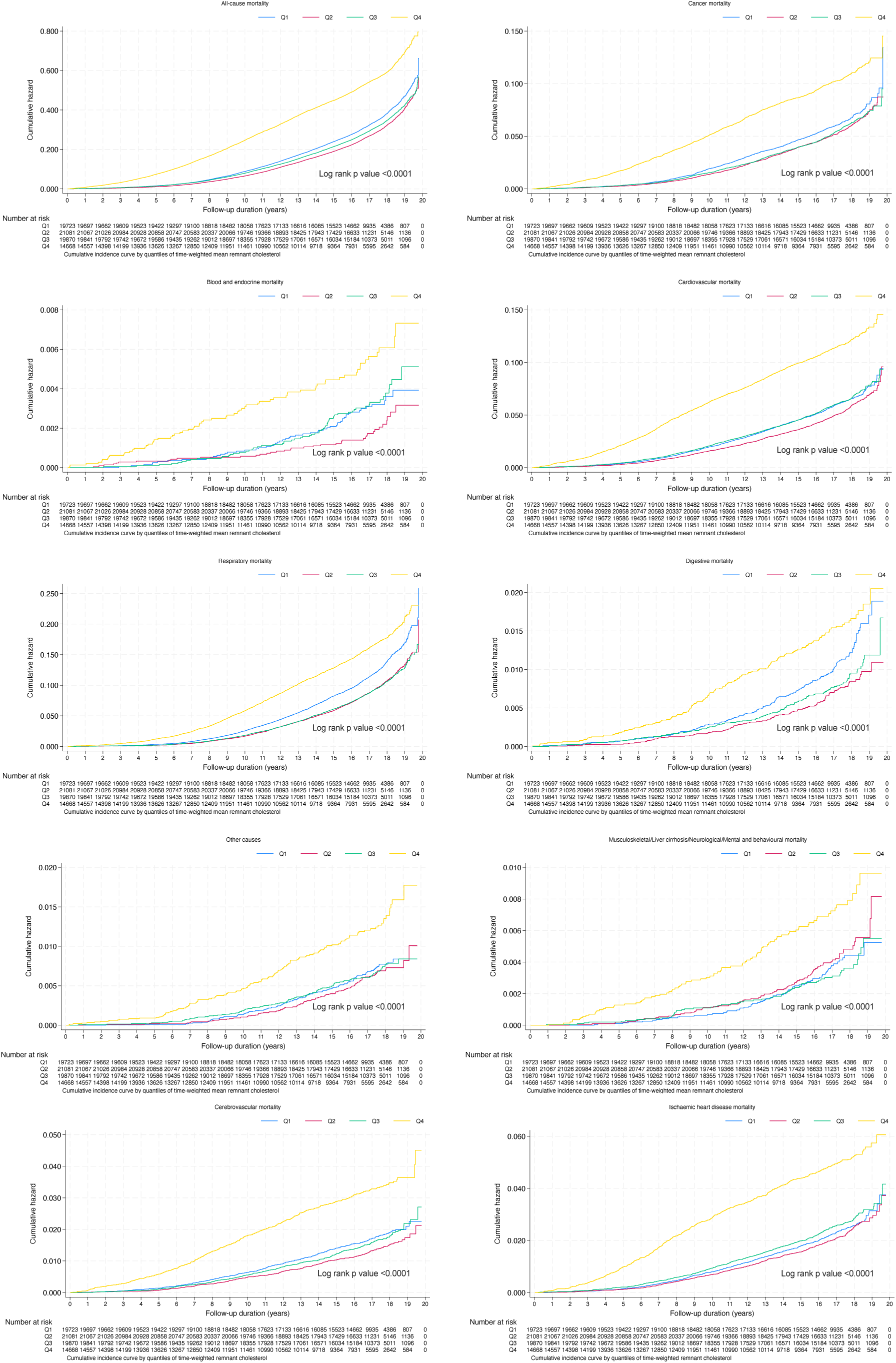
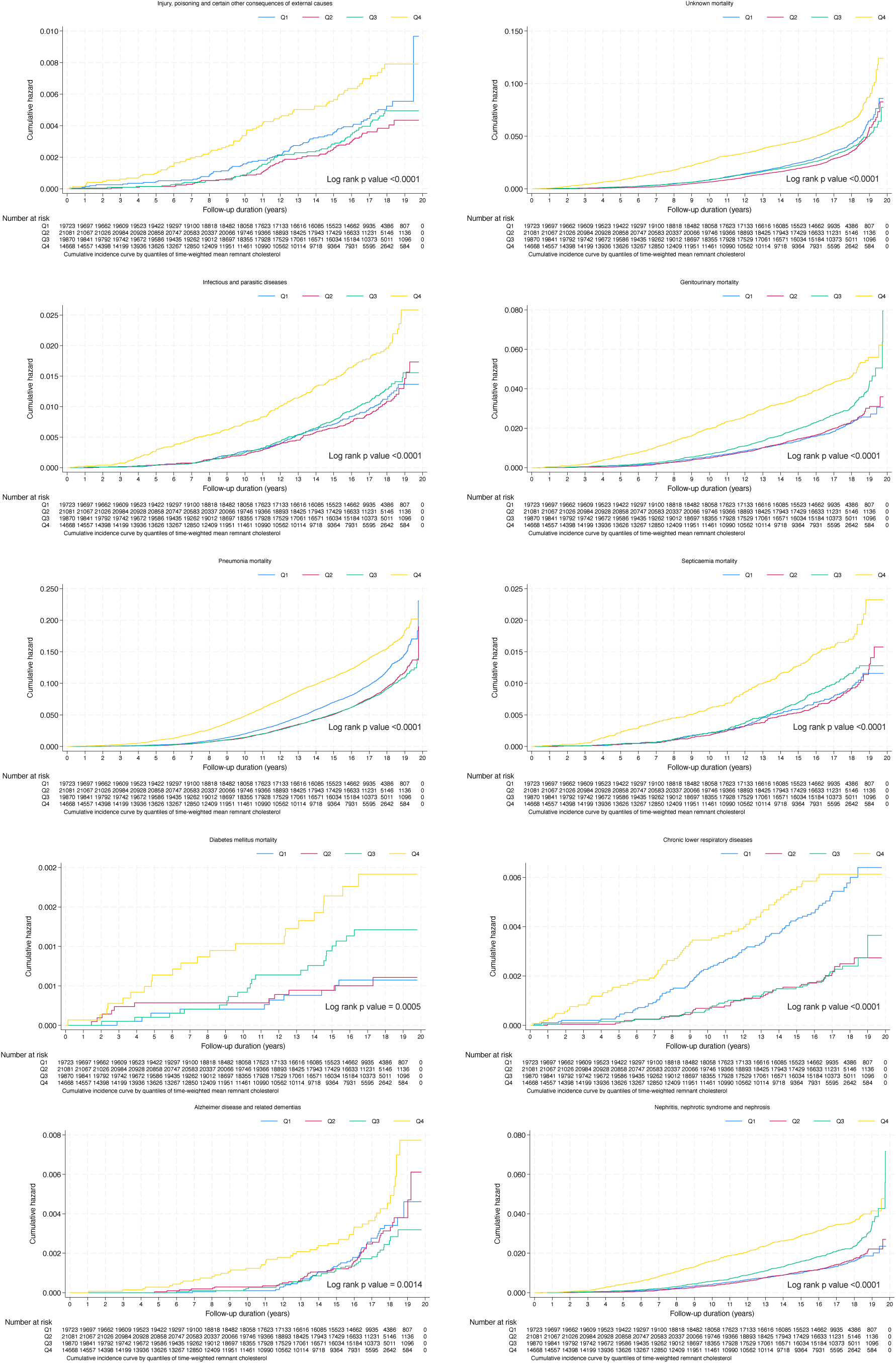
Cumulative incidence curve for cause-specific mortality by quantiles of remnant cholesterol.

**Figure 3.**
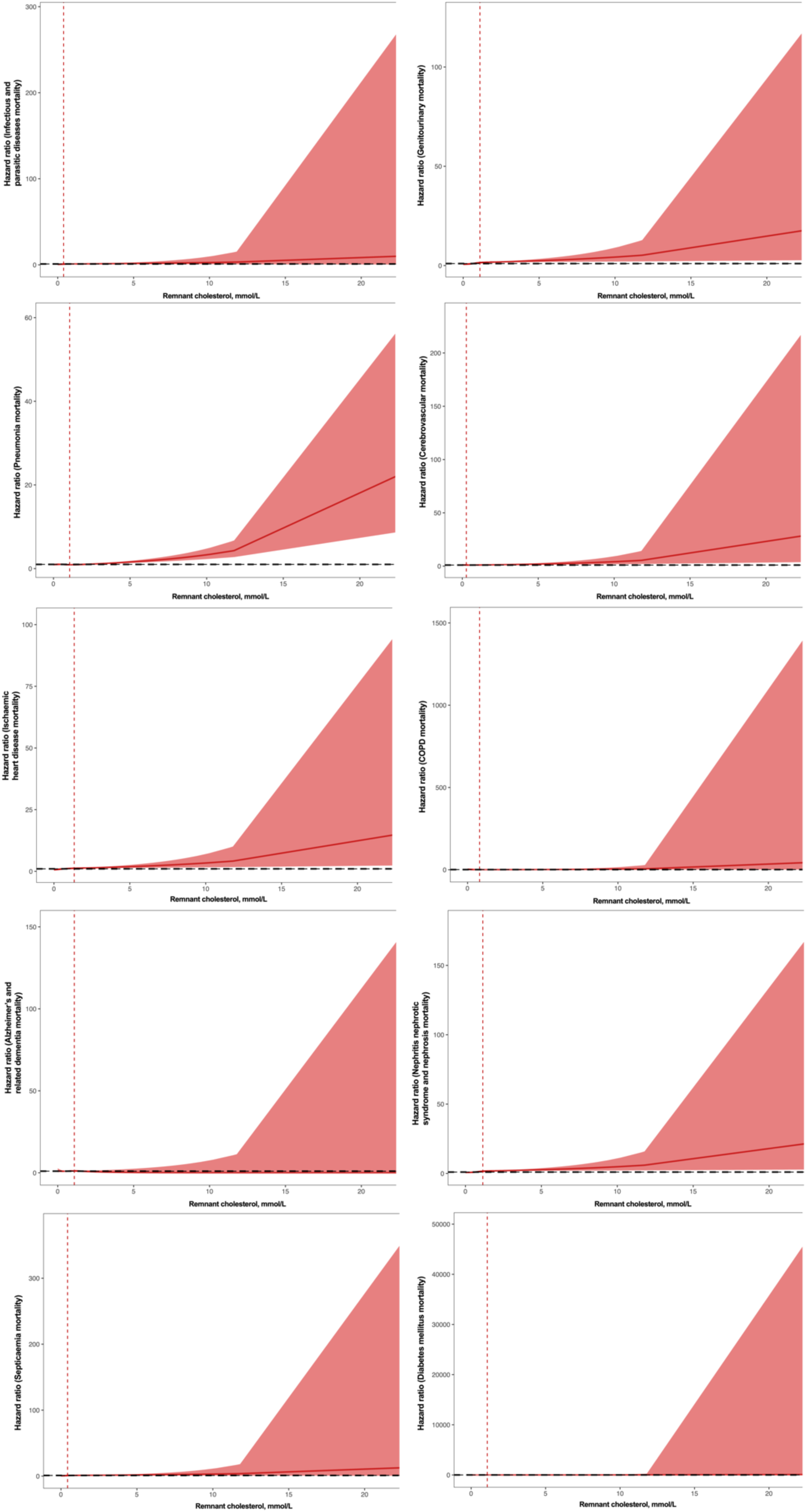
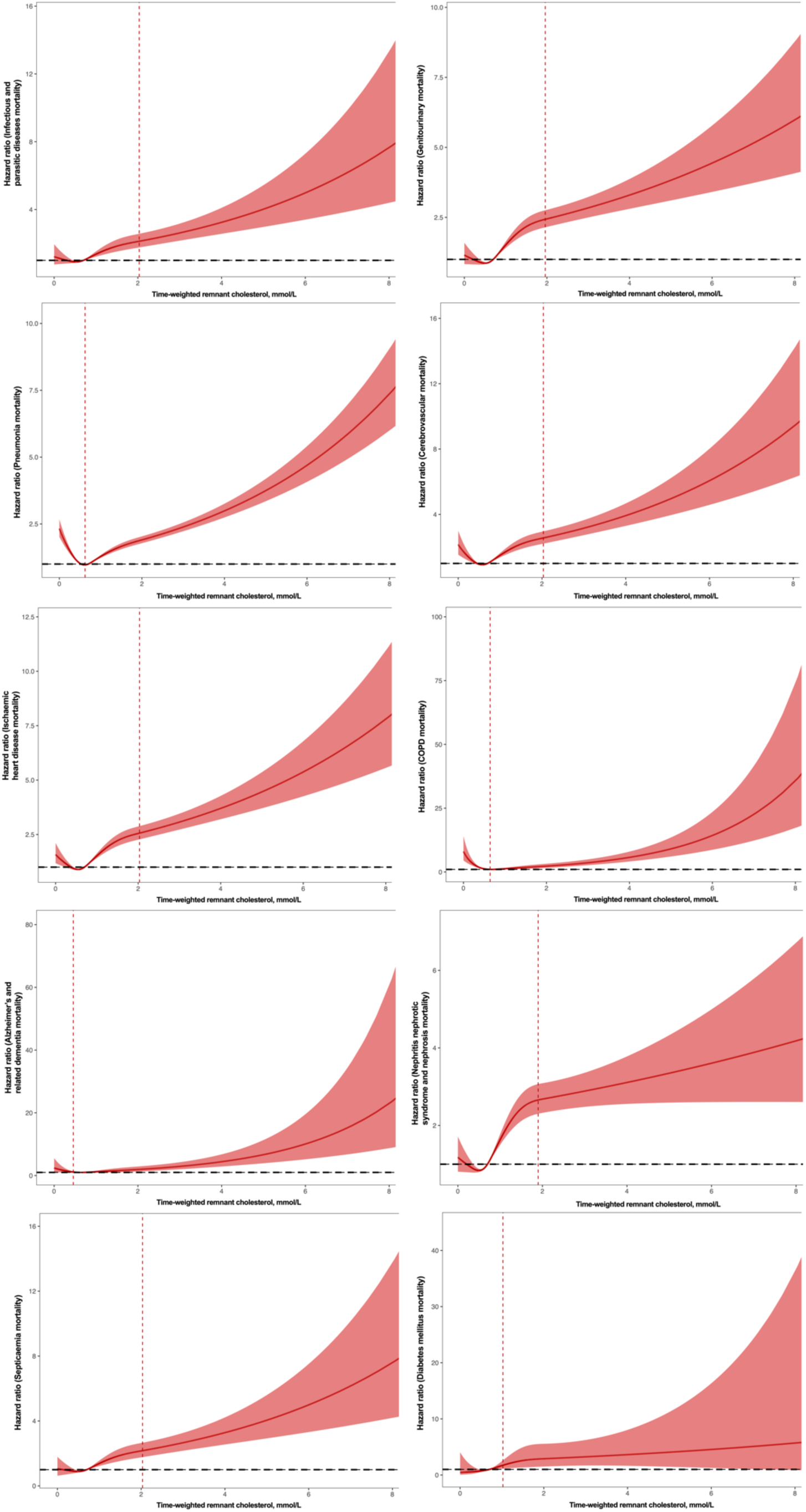
Restricted cubic spline for time-weighted remnant cholesterol predicting cause-related mortality on continuous scales.

### Subgroup analysis

Subgroup analysis **(Table 3)** showed that time-weighted RC associated with all-cause mortality, cardiovascular mortality, cancer mortality, and respiratory mortality within all subgroups of age, and statins and fibrates. Within the acute coronary syndrome subgroup, time-weighted RC also associated with all-cause mortality, and cardiovascular mortality, except for respiratory mortality.

**Table 3A.** Subgroup analysis to investigate the relationship between time-weighted remnant cholesterol and cause-specific mortality within different subgroups. * for p≤ 0.05, ** for p ≤ 0.01, *** for p ≤ 0.001, RC: remnant cholesterol, mmol/L, HR: hazard ratio, CI: confidence interval.

|  |  | All-cause mortality |  | Cancer-related mortality |  | Cardiovascular-related mortality |  | Ischaemic heart disease mortality |  | Respiratory-related mortality |  | Pneumonia-related mortality |  | Chronic lower respiratory mortality |  | Gastrointestinal-related mortality |  |
| --- | --- | --- | --- | --- | --- | --- | --- | --- | --- | --- | --- | --- | --- | --- | --- | --- | --- |
| Subgroup | Count(%) | Time-weighted RC HR[95%CI];p-value | P for interaction | Time-weighted RC HR[95%CI];p-value | P for interaction | HR[95%CI];p-value | P for interaction | Time-weighted RC HR[95%CI];p-value | P for interaction | Time-weighted RC HR[95%CI];p-value | P for interaction | Time-weighted RC HR[95%CI];p-value | P for interaction | Time-weighted RC HR[95%CI];p-value | P for interaction | Time-weighted RC HR[95%CI];p-value | P for interaction |
| Gender |  |  |  |  |  |  |  |  |  |  |  |  |  |  |  |  |  |
| Female | 45437(60.3%) | 1.41(1.38-1.43);<0.001** | <0.001** | 1.39(1.33-1.44);<0.001** | 0.673 | 1.47(1.42-1.51);<0.001** | 0.005* | 1.42(1.35-1.5);<0.001** | 0.864 | 1.39(1.35-1.43);<0.001** | <0.001** | 1.38(1.34-1.43);<0.001** | <0.001** | 1.58(1.34-1.86);<0.001** | 0.351 | 1.31(1.19-1.46); <0.001** | 0.156 |
| Male | 29905(39.7%) | 1.32(1.3-1.34);<0.001** |  | 1.37(1.32-1.42];<0.001** |  | 1.36(1.30-1.41);<0.001** |  | 1.4(1.33-1.48);<0.001** |  | 1.26(1.22-1.30);<0.001** |  | 1.24(1.19-1.28);<0.001** |  | 1.44(1.28-1.61);<0.001** |  | 1.17(1.02-1.33);0.023* |  |
| Age |  |  |  |  |  |  |  |  |  |  |  |  |  |  |  |  |  |
| ≥65, years | 32825(43.6%) | 1.25(1.24-1.27);<0.001** | <0.001** | 1.28(1.24-1.32);<0.001** | <0.001** | 1.30(1.27-1.34);<0.001** | <0.001** | 1.29(1.24-1.35);<0.001** | <0.001** | 1.22(1.19-1.25);<0.001** | <0.001** | 1.21(1.18-1.24);<0.001** | <0.001** | 1.33(1.2-1.48);<0.001** | 0.01* | 1.15(1.05-1.26);0.002* | 0.162 |
| <65, years | 42517(56.4%) | 1.46(1.42-1.50);0.001** |  | 1.44(1.38-1.52);<0.001** |  | 1.50(1.42-1.59);<0.001** |  | 1.55(1.42-1.69);<0.001** |  | 1.41(1.34-1.5);<0.001** |  | 1.38(1.3-1.48);<0.001** |  | 1.79(1.46-2.19);<0.001** |  | 1.32(1.10-1.58);0.002* |  |
| Dyslipidaemia |  |  |  |  |  |  |  |  |  |  |  |  |  |  |  |  |  |
| Yes | 1288(1.7%) | 1.56(1.44-1.69);<0.001** | <0.001** | 1.69(1.42-2.02);<0.001** | 0.003 | 1.41(1.14-1.72);0.001 | 0.790 | 1.55(1.2-2.01);0.001 | 0.368 | 1.57(1.34-1.83);<0.001** | 0.01* | 1.58(1.34-1.87);<0.001** | 0.009* | 1.37(0.57-3.32);0.483 | 0.909 | 1.53(0.99-2.36);0.053* | 0.257 |
| No | 74054(98.3%) | 1.37(1.35-1.38);<0.001** |  | 1.38(1.34-1.41);<0.001** |  | 1.42(1.39-1.46);<0.001** |  | 1.42(1.37-1.47);<0.001** |  | 1.33(1.30-1.36);<0.001** |  | 1.31(1.28-1.34);<0.001** |  | 1.5(1.36-1.65);<0.001** |  | 1.25(1.15-1.35);<0.001** |  |
| Diabetes mellitus |  |  |  |  |  |  |  |  |  |  |  |  |  |  |  |  |  |
| Yes | 7889(10.5%) | 1.67(1.62-1.73);<0.001** | <0.001** | 1.72(1.60-1.86);<0.001** | <0.001** | 1.74(1.63-1.85);<0.001** | <0.001** | 1.63(1.48-1.8);<0.001** | 0.004 | 1.62(1.51-1.74);<0.001** | <0.001** | 1.64(1.52-1.77);<0.001** | <0.001** | 1.44(0.88-2.37);0.15 | 0.874 | 1.60(1.3-1.97);<0.001** | 0.013* |
| No | 67453(89.5%) | 1.35(1.34-1.37);<0.001** |  | 1.36(1.32-1.40);<0.001** |  | 1.40(1.36-1.44);<0.001** |  | 1.42(1.36-1.48);<0.001** |  | 1.32(1.29-1.35);<0.001** |  | 1.3(1.27-1.33);<0.001** |  | 1.5(1.36-1.65);<0.001** |  | 1.23(1.12-1.34);<0.001** |  |

| Hypertension |  |  |  |  |  |  |  |  |  |  |  |  |  |  |  |  |  |
| --- | --- | --- | --- | --- | --- | --- | --- | --- | --- | --- | --- | --- | --- | --- | --- | --- | --- |
| Yes | 39026(51.8%) | 1.46(1.44-1.499);<0.001** | <0.001** | 1.48(1.43-1.53);<0.001** | <0.001** | 1.49(1.44-1.54);<0.001** | 0.029* | 1.48(1.41-1.56);<0.001** | 0.330 | 1.44(1.40-1.48);<0.001** | <0.001** | 1.42(1.37-1.46);<0.001** | <0.001** | 1.6(1.41-1.82);<0.001** | 0.301 | 1.30(1.16-1.45);<0.001** | 0.717 |
| No | 36316(48.2%) | 1.33(1.30-1.35);<0.001** |  | 1.33(1.28-1.38);<0.001** |  | 1.41(1.36-1.47);<0.001** |  | 1.42(1.34-1.51);<0.001** |  | 1.28(1.23-1.32);<0.001** |  | 1.27(1.23-1.32);<0.001** |  | 1.44(1.24-1.66);<0.001** |  | 1.26(1.12-1.43);<0.001** |  |
| Acute coronary artery syndrome |  |  |  |  |  |  |  |  |  |  |  |  |  |  |  |  |  |
| Yes | 173(0.2%) | 1.79(1.44-2.22);<0.001** | <0.001** | 1.60(0.90-2.85);0.109 | 0.262 | 2.17(1.57-3.01);<0.001** | <0.001** | 2.23(1.39-3.58);0.001* | 0.015* | 1.35(0.66-2.79);0.411 | 0.782 | 1.4(0.7-2.82);0.346 | 0.64 | 1.57(1.27-1.95);<0.001** | 0.356 | 9.05(0.41-200.43);0.163 | 0.473 |
| No | 75169(99.8%) | 1.37(1.35-1.38);<0.001** |  | 1.38(1.34-1.42);<0.001** |  | 1.42(1.38-1.45);<0.001** |  | 1.42(1.37-1.47);<0.001** |  | 1.33(1.30-1.36);<0.001** |  | 1.31(1.28-1.35);<0.001** |  | 1.5(1.36-1.64);<0.001** |  | 1.25(1.15-1.36);<0.001** |  |
| Heart failure |  |  |  |  |  |  |  |  |  |  |  |  |  |  |  |  |  |
| Yes | 2932(3.9%) | 1.24(1.20-1.29);<0.001** | <0.001** | 1.29(1.17-1.42);<0.001** | 0.207 | 1.30(1.22-1.39);<0.001** | 0.121 | 1.27(1.15-1.41);<0.001** | 0.163 | 1.31(1.04-1.22);0.004 | <0.001** | 1.15(1.06-1.25);0.001 | 0.005* | 0.85(0.46-1.58);0.607 | 0.069 | 1.21(0.96-1.51);0.1 | 0.943 |
| No | 72410(96.1%) | 1.37(1.35-1.39);<0.001** |  | 1.38(1.35-1.42);<0.001** |  | 1.41(1.37-1.45);<0.001** |  | 1.42(1.36-1.47);<0.001** |  | 1.34(1.31-1.37);<0.001** |  | 1.32(1.29-1.35);<0.001** |  | 1.54(1.4-1.69);<0.001** |  | 1.24(1.13-1.35);<0.001** |  |
| Atrial fibrillation |  |  |  |  |  |  |  |  |  |  |  |  |  |  |  |  |  |
| Yes | 446(0.6%) | 1.44(1.32-1.57);<0.001** | 0.056 | 1.61(1.26-2.04);0.001** | 0.113 | 1.50(1.30-1.74);<0.001** | 0.148 | 1.91(1.57-2.32);<0.001** | 0.001* | 1.36(1.13-1.63);0.001 | 0.641 | 1.3(1.05-1.62);0.016 | 0.907 | 2.13(1.31-3.46);0.002 | 0.061 | 1.35(0.79-2.32);0.276 | 0.479 |
| No | 74896(99.4%) | 1.37(1.35-1.38);<0.001** |  | 1.38(1.34-1.42);<0.001** |  | 1.42(1.38-1.45);<0.001** |  | 1.41(1.36-1.46);<0.001** |  | 1.33(1.30-1.36);<0.001** |  | 1.32(1.28-1.35);<0.001** |  | 1.48(1.34-1.63);<0.001** |  | 1.25(1.15-1.36);<0.001** |  |
| Statins and fibrates |  |  |  |  |  |  |  |  |  |  |  |  |  |  |  |  |  |
| Yes | 8460(11.2%) | 1.39(1.34-1.43);<0.001** | 0.068 | 1.44(1.34-1.54);<0.001** | 0.065 | 1.35(1.27-1.44);<0.001** | 0.247 | 1.37(1.26-1.5);<0.001** | 0.645 | 1.33(1.25-1.42);<0.001** | 0.812 | 1.33(1.24-1.43);<0.001** | 0.619 | 1.39(1.01-1.92);0.046 | 0.676 | 1.46(1.25-1.71);<0.001** | 0.012* |
| No | 66882(88.8%) | 1.36(1.3501.38);<0.001** |  | 1.37(1.33-1.41);<0.001** |  | 1.43(1.39-1.47);<0.001** |  | 1.43(1.37-1.49);<0.001** |  | 1.33(1.29-1.36);<0.001** |  | 1.31(1.28-1.34);<0.001** |  | 1.51(1.37-1.66);<0.001** |  | 1.20(1.09-1.32);<0.001** |  |

| Nitrates |  |  |  |  |  |  |  |  |  |  |  |  |  |  |  |  |  |
| --- | --- | --- | --- | --- | --- | --- | --- | --- | --- | --- | --- | --- | --- | --- | --- | --- | --- |
| Yes | 6148(8.2%) | 1.37(1.33-1.41);<0.001** | 0.130 | 1.38(1.28-1.48);<0.001** | 0.663 | 1.36(1.29-1.44);<0.001** | 0.453 | 1.37(1.27-1.48);<0.001** | 0.895 | 1.33(1.26-1.41);<0.001** | 0.463 | 1.32(1.24-1.4);<0.001** | 0.667 | 1.28(0.99-1.64);0.061 | 0.235 | 1.29(1.07-1.57);0.009* | 0.616 |
| No | 60194(91.8%) | 1.36(1.34-1.37);<0.001** |  | 1.37(1.34-1.41);<0.001** |  | 1.41(1.37-1.45);<0.001** |  | 1.4(1.34-1.47);<0.001** |  | 1.32(1.29-1.35);<0.001** |  | 1.31(1.27-1.34);<0.001** |  | 1.53(1.38-1.69);<0.001** |  | 1.24(1.13-1.35);<0.001** |  |

**Table 3B.**
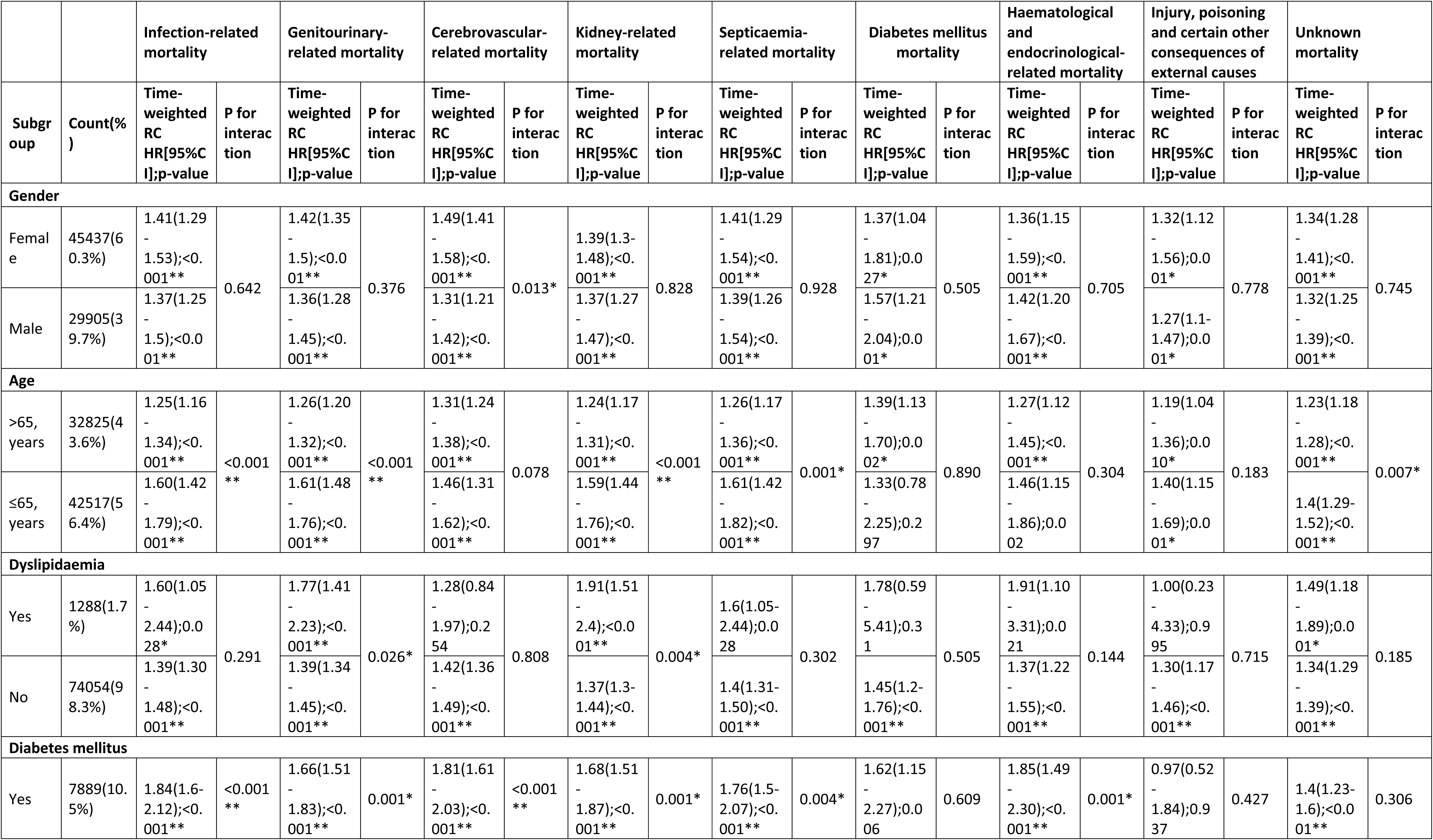
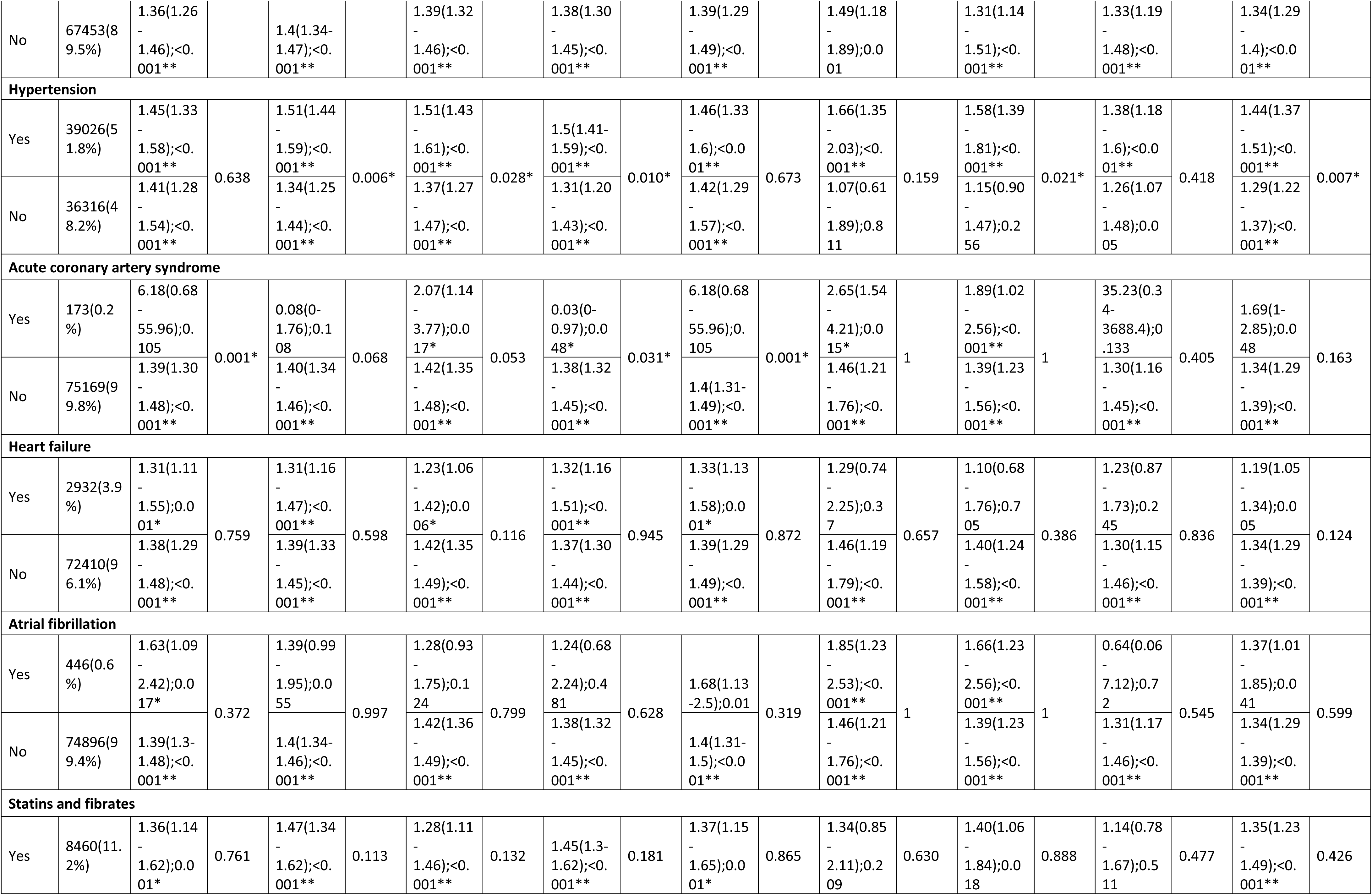
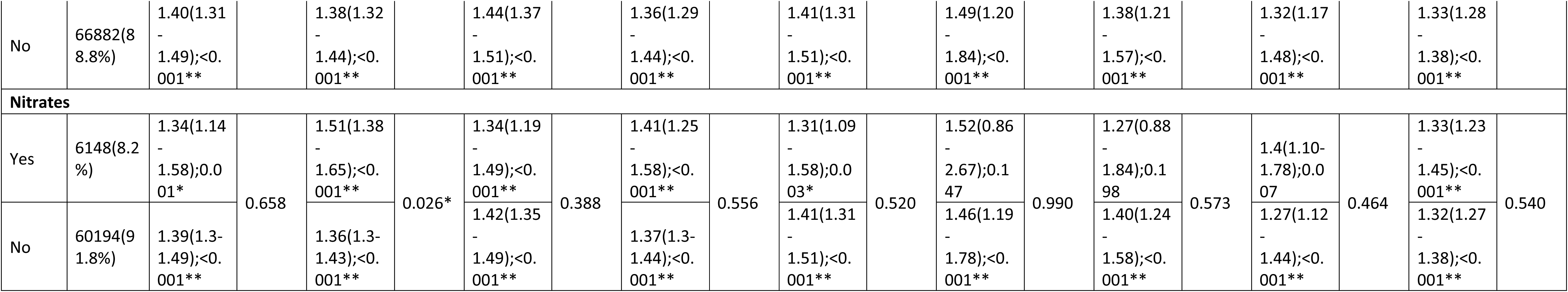
Subgroup analysis to investigate the relationship between time-weighted remnant cholesterol and cause-specific mortality within different subgroups. * for p≤ 0.05, ** for p ≤ 0.01, *** for p ≤ 0.001, RC: remnant cholesterol, HR: hazard ratio, CI: confidence interval.

Specifically, time-weighted RC associated with cardiovascular mortality in both aged ≥65 (HR: 1.30; CI: 1.27-1.34; p<0.001) and aged <65 (HR: 1.50; CI: 1.42-1.59; p<0.001) subgroups. Similar observations were found in individuals that had acute coronary syndrome (HR: 2.23; CI: 1.39-3.58; p=0.001) and without acute coronary syndrome (1.42(1.37-1.47); p<0.001), individuals that take statins and fibrates (HR: 1.35; CI: 1.27-1.44; p<0.001), and in individuals that did not receive statins nor fibrates (HR: 1.43; CI: 1.39-1.47; p<0.001).

## Discussion

In this population-based cohort study with over 15 years of follow-up, time-weighted RC was associated with all-cause mortality amongst patients attending the family medicine clinic. Furthermore, time-weighted RC was associated with cardiovascular-related mortality, cancer-related mortality, and respiratory-related mortality, and other mortality outcomes apart from gastrointestinal-related mortality, diabetes mellitus mortality and unknown mortality. To the best of our knowledge, the present study is the first to demonstrate the use of time-weighted RC to predict cause-specific mortalities in the general population.

### Comparison with previous studies

The association with time weighted RC and CVD-related mortality obtained in this cohort study is comparable to previous studies. Elevated levels of RC were previously shown to be associated with increased risks of peripheral artery disease (10), ischaemic heart disease (11–15), and ischaemic stroke (16), all of which encompass conditions with high mortality. Elevated RC have previously been associated with poor outcomes in individuals who have suffered from myocardial infarction with non-obstructive coronary arteries, which commonly occurs in younger patients with fewer comorbidities (17). Wadstrom *et al.* conducted a prospective study and found that individuals with ≥1 mmol/L of RC had two-fold rate of cardiovascular mortality, amongst other causes of mortality (18). Moreover, *Tian et al.* further supported these findings with demonstrating an association with high RC levels and cardiovascular-related mortality in a prospective study (19). Additionally, within the same study, low levels of RC were associated with increased risk of cerebrovascular mortality, which is a novel finding. Our results similarly demonstrated such an association between time-weighted RC and cerebrovascular mortality.

Non-cardiovascular diseases known to be associated with increased mortality, such as cancer, severe respiratory diseases, inflammatory diseases, are associated with reduced concentrations of LDL and total cholesterol (20, 21). However, our findings on cancer and respiratory mortality did not support these findings.

To the best of our knowledge, we are the first to demonstrate an association with cancer mortality. Other studies seem to have negative or contrasting results. In the study of Wadstrom *et al.*, individuals with high RC levels did not show any correlations to cancer mortality (18). Similarly, Bonfiglio *et al.* found no increased risk for cancer mortality in individuals with increased RC within their study (22). Interestingly, *Tian et al.* demonstrated a reduced risk of overall cancer mortality and specific types of cancer mortality in individuals with high RC (19). Perhaps, future studies could explore the relationship between RC and cancer mortality in more depth by cancer types.

Our findings surrounding the association with respiratory-related mortality is novel and difficult to explain. In the sole comparable study, no association was found between increased levels of RC and respiratory mortality (18). The differences in results may be attributable to the difference in demographics and epidemiology within the populations across studies. Nonetheless, further research is warranted to ascertain the relationship between elevated levels of RC and other causes of mortality.

### Mechanism of action

RC, like LDL-C particles, may accumulate within the arterial wall and cause atherosclerosis due to its cholesterol composition (23). RC seems to directly correlate with increased levels of triglyceride, making triglycerides a good indicator for RC levels (24, 25). However, the atherosclerotic effect of RC seems to act irrespective of blood triglyceride levels; a study showed an association between elevated levels of RC and increased risk of myocardial infarction within a group of individuals with high triglyceride (12).

Another established causative link to atherosclerosis is apolipoprotein (ApoB) levels (26). A large cohort study found the risk of myocardial infarction was most associated to the level of ApoB-containing lipoproteins, independent of the lipid content or type of lipoprotein (27). About one third of the cholesterol carried in ApoB is transported through remnant particles in non-fasting states (28). However, a study in cardiovascular-free individuals found an association between elevated RC levels and atherosclerotic cardiovascular diseases, independent of ApoB levels (29). This may indicate that RC may have its own mechanism of action at causing atherosclerosis, unrelated to ApoB, which may be worth investigating.

RC does not only potentiate atherosclerosis, it has also been associated with causing low-grade inflammation and oxidative stress (30, 31), as opposed to LDL-C which only causes ischaemic heart disease (13). Through complex physiological processes, RC is believed to contribute to driving obesity and cancer progression (32–34). However, the role of RC is less clear in conditions outside of atherosclerotic cardiovascular diseases.

### Clinical relevance

Despite efforts of lowering of LDL-C through statin therapy, significant cardiovascular risk remains in individuals treated with statin, even when low LDL-C levels have been achieved (35, 36). RC is an emerging risk factor for atherosclerotic cardiovascular disease, which could prove to be an effective potential pharmacological target at reducing mortality. Some drugs in development are showing promising results in lowering RC and triglycerides (37–39). Our findings support that RC lowering drugs might improve cardiovascular disease outcomes. However, further prospective studies should be done to confirm the causation relationship with other causes of mortality in individuals with elevated RC, to evaluate whether RC-lowering treatment has a role outside of preventing cardiovascular mortality.

### Limitations

Several limitations are present in this study which should be acknowledged. Firstly, the derived populations may differ from other populations due to both comorbidities and demographic differences. The model should be externally validated using patient data from other regions. Secondly, given the retrospective nature of this study, the residual or unmeasured confounders were not addressed. Besides, the observational study was also subjected to bias secondary to under-coding and documentation errors. Some covariates associated with mortality, such as smoking, alcohol uses, and obesity were not included in this study. Last but not least, this retrospective study could only demonstrate the association but not the causation relationship between RC and mortality.

## Conclusions

Time-weighted RC was associated with all-cause mortality and cause-specific mortality outcomes amongst the general population. Optimal RC control may not only help prevent cardiovascular mortality but mortality due to other causes.

## Data Availability

The data that support the findings of this study were provided by the Hong Kong Hospital Authority, but restrictions apply to the availability of these data, which were used under license for the current study, and so are not publicly available. Data are however available from the authors upon reasonable request and with permission of Hong Kong Hospital Authority.

## Author contributions

Lifang Li, Vanessa Hou Cheng Chou, Oscar Hou In Chou, Jiandong Zhou: Conception of study, preparation of figures, study design, data contribution, statistical analysis, data interpretation, manuscript drafting, and critical revision of the manuscript.

Sakshi Roy, Jeffrey Shi Kai Chan, Wing Tak Wong, Tong Liu: Data interpretation, literature search, data collection, manuscript drafting.

Gregory Lip, Bernard Man Yung Cheung: Literature search, critical revision of the manuscript

Gary Tse, Jiandong Zhou: Conception of study and literature search, study design, data collection, and critical revision of manuscript, study supervision.

## Ethical approval and consent to participate

This study was approved by the Institutional Review Board of the University of Hong Kong/ Hospital Authority Hong Kong West Cluster (Reference No. UW 20-250).

## Acknowledgements

All the authors and colleagues from the Hospital Authority for providing de-identified clinical data are equally thanked for their contributions to this research. Special thanks to the support of the National Natural Science Foundation of China (81970270, 82170327 to TL) and the Tianjin Key Medical Discipline (Specialty) Construction Project (TJYXZDXK-029A). Structural graphical abstract and Figure 1 are created with BioRender.com.

## Guarantor Statement

All authors approved the final version of the manuscript. GT is the guarantor of this work and, as such, had full access to all the data in the study and takes responsibility for the integrity of the data and the accuracy of the data analysis.

