## Supplementary Appendix for "The association between time-weighted remnant cholesterol and cardiovascular and non-cardiovascular mortality: A population-based cohort study"

**Supplementary Table 1. International Classification of Diseases Ninth Version (ICD-9) codes for baseline comorbidities.**

| | |
|---|---|
| Diabetes mellitus | 250 250.01 250.02 250.03 250.1 250.11 250.12 250.13 250.2 250.21 250.22 250.23 250.3 250.31 250.32 250.33 250.4 250.41 250.42 250.43 250.5 250.51 250.52 250.53 250.6 250.61 250.62 250.63 250.7 250.71 250.72 250.73 250.8 250.81 250.82 250.83 250.9 250.91 250.92 250.93 |
| Renal diseases | 582 582 582.1 582.2 582.4 582.8 582.81 582.89 582.9 583 583 583.1 583.2 583.4 583.6 583.7 585 585.1 585.2 585.3 585.4 585.5 585.6 585.9 586 588 588 588.1 588.8 588.81 588.89 588.9 |
| Cerebrovascular disease | 430 431 432 432.1 432.9 433 433.01 433.1 433.11 433.2 433.21 433.3 433.31 433.8 433.81 433.9 433.91 434 434.01 434.1 434.11 434.9 434.91 435 435.1 435.2 435.3 435.8 435.9 436 437 437.1 437.2 437.3 437.4 437.5 437.6 437.7 437.8 437.9 438 438.1 438.11 438.12 438.13 438.14 438.19 438.2 438.21 438.22 438.3 438.31 438.32 438.4 438.41 438.42 438.5 438.51 438.52 438.53 438.6 438.7 438.8 438.81 438.82 438.83 438.84 438.85 438.89 438.9 |
| Acute myocardial infarction | 410 410 410 410.01 410.02 410.1 410.1 410.11 410.12 410.2 410.2 410.21 410.22 410.3 410.3 410.31 410.32 410.4 410.4 410.41 410.42 410.5 410.5 410.51 410.52 410.6 410.6 410.61 410.62 410.7 410.7 410.71 410.72 410.8 410.8 410.81 410.82 410.9 410.9 410.91 410.92 412 |
| Hypertension | 401 401.1 401.9 402 402.01 402.1 402.11 402.9 402.91 403 403.01 403.1 403.11 403.9 403.91 404 404.01 404.02 404.03 404.1 404.11 404.12 404.13 404.9 404.91 404.92 404.93 405 405.01 405.09 405.1 405.11 405.19 405.9 405.91 405.99 437.2 |
| Heart failure | 428 428 428.1 428.2 428.2 428.21 428.22 428.23 428.3 428.3 428.31 428.32 428.33 428.4 428.4 428.41 428.42 428.43 428.9 398.91 402.01 402.11 402.91 404.01 404.03 404.11 404.13 404.91 404.93 |
| Atrial fibrillation | 427.31 429.4 |
| Liver diseases | 456 456.1 456.2 572.2 572.3 572.4 572.8 571.4 571.5 571.6 |
| COPD | 490 491 492 493 494 495 496 491.1 491.2 491.21 491.22 491.8 491.9 492.8 493.01 493.02 493.1 493.11 493.12 493.2 493.21 493.22 493.8 493.81 493.82 493.9 493.91 493.92 494.1 495.1 495.2 495.3 495.4 495.5 495.6 495.7 495.8 495.9 |
| Ischaemic heart diseases | 410.01 410.02 410.1 410.11 410.12 410.2 410.21 410.22 410.3 410.31 410.32 410.4 410.41 410.42 410.5 410.51 410.52 410.6 410.61 410.62 410.7 410.71 410.72 410.8 410.81 410.82 410.9 410.91 410.92 411 411.1 411.8 411.81 411.89 413 413.1 413.9 414 414.01 414.02 414.03 414.04 414.05 414.06 414.07 414.1 414.11 414.12 414.19 414.2 414.3 414.4 414.8 414.9 410 412 |
| Cancer | 140 140.1 140.3 140.4 140.5 140.6 140.8 140.9 141 141.1 141.2 141.3 141.4 141.5 141.6 141.8 141.9 142 142.1 142.2 142.8 142.9 143 143.1 143.8 143.9 144 144.1 144.8 144.9 145 145.1 145.2 145.3 145.4 145.5 145.6 145.8 145.9 146 146.1 146.2 146.3 146.4 146.5 146.6 146.7 146.8 146.9 147 147.1 147.2 147.3 147.8 147.9 148 148.1 148.2 148.3 148.8 148.9 149 149.1 149.8 149.9 150 |

| | | | | | | | | | | |
|---|---|---|---|---|---|---|---|---|---|---|
| 150.1 | 150.2 | 150.3 | 150.4 | 150.5 | 150.8 | 150.9 | 151 | 151.1 | 151.2 | 151.3 |
| 151.4 | 151.5 | 151.6 | 151.8 | 151.9 | 152 | 152.1 | 152.2 | 152.3 | 152.8 | 152.9 |
| 153 | 153.1 | 153.2 | 153.3 | 153.4 | 153.5 | 153.6 | 153.7 | 153.8 | 153.9 | 154 |
| 154.1 | 154.2 | 154.3 | 154.8 | 155 | 155.1 | 155.2 | 156 | 156.1 | 156.2 | 156.8 |
| 156.9 | 157 | 157.1 | 157.2 | 157.3 | 157.4 | 157.8 | 157.9 | 158 | 158.8 | 158.9 |
| 159 | 159.1 | 159.8 | 159.9 | 160 | 160.1 | 160.2 | 160.3 | 160.4 | 160.5 | 160.8 |
| 160.9 | 161 | 161.1 | 161.2 | 161.3 | 161.8 | 161.9 | 162 | 162.2 | 162.3 | 162.4 |
| 162.5 | 162.8 | 162.9 | 163 | 163.1 | 163.8 | 163.9 | 164 | 164.1 | 164.2 | 164.3 |
| 164.8 | 164.9 | 165 | 165.8 | 165.9 | 170 | 170.1 | 170.2 | 170.3 | 170.4 | 170.5 |
| 170.6 | 170.7 | 170.8 | 170.9 | 171 | 171.2 | 171.3 | 171.4 | 171.5 | 171.6 | 171.7 |
| 171.8 | 171.9 | 172 | 172.1 | 172.2 | 172.3 | 172.4 | 172.5 | 172.6 | 172.7 | 172.8 |
| 172.9 | and | 173 | 173.01 | 173.02 | 173.09 | 173.1 | 173.11 | 173.12 | 173.19 | 173.2 |
| 173.21 | 173.22 | 173.29 | 173.3 | 173.31 | 173.32 | 173.39 | 173.4 | 173.41 | 173.42 | 173.49 |
| 173.5 | 173.51 | 173.52 | 173.59 | 173.6 | 173.61 | 173.62 | 173.69 | 173.7 | 173.71 | 173.72 |
| 173.79 | 173.8 | 173.81 | 173.82 | 173.89 | 173.9 | 173.91 | 173.92 | 173.99 | 174 | 174.1 |
| 174.2 | 174.3 | 174.4 | 174.5 | 174.6 | 174.8 | 174.9 | 175 | 175.9 | 176 | 176.1 |
| 176.2 | 176.3 | 176.4 | 176.5 | 176.8 | 176.9 | 179 | 180 | 180.1 | 180.8 | 180.9 |
| 181 | 182 | 182.1 | 182.8 | 183 | 183.2 | 183.3 | 183.4 | 183.5 | 183.8 | 183.9 |
| 184 | 184.1 | 184.2 | 184.3 | 184.4 | 184.8 | 184.9 | 185 | 186 | 186.9 | 187 |
| 187.1 | 187.2 | 187.3 | 187.4 | 187.5 | 187.6 | 187.7 | 187.8 | 187.9 | 188 | 188.1 |
| 188.2 | 188.3 | 188.4 | 188.5 | 188.6 | 188.7 | 188.8 | 188.9 | 189 | 189.1 | 189.2 |
| 189.3 | 189.4 | 189.8 | 189.9 | 190 | 190.1 | 190.2 | 190.3 | 190.4 | 190.5 | 190.6 |
| 190.7 | 190.8 | 190.9 | 191 | 191.1 | 191.2 | 191.3 | 191.4 | 191.5 | 191.6 | 191.7 |
| 191.8 | 191.9 | 192 | 192.1 | 192.2 | 192.3 | 192.8 | 192.9 | 193 | 194 | 194.1 |
| 194.3 | 194.4 | 194.5 | 194.6 | 194.8 | 194.9 | 195 | 195.1 | 195.2 | 195.3 | 195.4 |
| 195.5 | 195.8 | 200 | 200.01 | 200.02 | 200.03 | 200.04 | 200.05 | 200.06 | 200.07 | 200.08 |
| 200.1 | 200.11 | 200.12 | 200.13 | 200.14 | 200.15 | 200.16 | 200.17 | 200.18 | 200.2 | 200.21 |
| 200.22 | 200.23 | 200.24 | 200.25 | 200.26 | 200.27 | 200.28 | 200.3 | 200.31 | 200.32 | 200.33 |
| 200.34 | 200.35 | 200.36 | 200.37 | 200.38 | 200.4 | 200.41 | 200.42 | 200.43 | 200.44 | 200.45 |
| 200.46 | 200.47 | 200.48 | 200.5 | 200.51 | 200.52 | 200.53 | 200.54 | 200.55 | 200.56 | 200.57 |
| 200.58 | 200.6 | 200.61 | 200.62 | 200.63 | 200.64 | 200.65 | 200.66 | 200.67 | 200.68 | 200.7 |
| 200.71 | 200.72 | 200.73 | 200.74 | 200.75 | 200.76 | 200.77 | 200.78 | 200.8 | 200.81 | 200.82 |
| 200.83 | 200.84 | 200.85 | 200.86 | 200.87 | 200.88 | 201 | 201.01 | 201.02 | 201.03 | 201.04 |
| 201.05 | 201.06 | 201.07 | 201.08 | 201.1 | 201.11 | 201.12 | 201.13 | 201.14 | 201.15 | 201.16 |
| 201.17 | 201.18 | 201.2 | 201.21 | 201.22 | 201.23 | 201.24 | 201.25 | 201.26 | 201.27 | 201.28 |
| 201.4 | 201.41 | 201.42 | 201.43 | 201.44 | 201.45 | 201.46 | 201.47 | 201.48 | 201.5 | 201.51 |
| 201.52 | 201.53 | 201.54 | 201.55 | 201.56 | 201.57 | 201.58 | 201.6 | 201.61 | 201.62 | 201.63 |
| 201.64 | 201.65 | 201.66 | 201.67 | 201.68 | 201.7 | 201.71 | 201.72 | 201.73 | 201.74 | 201.75 |
| 201.76 | 201.77 | 201.78 | 201.9 | 201.91 | 201.92 | 201.93 | 201.94 | 201.95 | 201.96 | 201.97 |
| 201.98 | 202 | 202.01 | 202.02 | 202.03 | 202.04 | 202.05 | 202.06 | 202.07 | 202.08 | 202.1 |
| 202.11 | 202.12 | 202.13 | 202.14 | 202.15 | 202.16 | 202.17 | 202.18 | 202.2 | 202.21 | 202.22 |
| 202.23 | 202.24 | 202.25 | 202.26 | 202.27 | 202.28 | 202.3 | 202.31 | 202.32 | 202.33 | 202.34 |
| 202.35 | 202.36 | 202.37 | 202.38 | 202.4 | 202.41 | 202.42 | 202.43 | 202.44 | 202.45 | 202.46 |
| 202.47 | 202.48 | 202.5 | 202.51 | 202.52 | 202.53 | 202.54 | 202.55 | 202.56 | 202.57 | 202.58 |
| 202.6 | 202.61 | 202.62 | 202.63 | 202.64 | 202.65 | 202.66 | 202.67 | 202.68 | 202.7 | 202.71 |
| 202.72 | 202.73 | 202.74 | 202.75 | 202.76 | 202.77 | 202.78 | 202.8 | 202.81 | 202.82 | 202.83 |
| 202.84 | 202.85 | 202.86 | 202.87 | 202.88 | 202.9 | 202.91 | 202.92 | 202.93 | 202.94 | 202.95 |

| | | | | | | | | | | |
|---|---|---|---|---|---|---|---|---|---|---|
| 202.96 | 202.97 | 202.98 | 203 | 203.01 | 203.02 | 203.1 | 203.11 | 203.12 | 203.8 | 203.81 |
| 203.82 | 204 | 204.01 | 204.02 | 204.1 | 204.11 | 204.12 | 204.2 | 204.21 | 204.22 | 204.8 |
| 204.81 | 204.82 | 204.9 | 204.91 | 204.92 | 205 | 205.01 | 205.02 | 205.1 | 205.11 | 205.12 |
| 205.2 | 205.21 | 205.22 | 205.3 | 205.31 | 205.32 | 205.8 | 205.81 | 205.82 | 205.9 | 205.91 |
| 205.92 | 206 | 206.01 | 206.02 | 206.1 | 206.11 | 206.12 | 206.2 | 206.21 | 206.22 | 206.8 |
| 206.81 | 206.82 | 206.9 | 206.91 | 206.92 | 207 | 207.01 | 207.02 | 207.1 | 207.11 | 207.12 |
| 207.2 | 207.21 | 207.22 | 207.8 | 207.81 | 207.82 | 208 | 208.01 | 208.02 | 208.1 | 208.11 |
| 208.12 | 208.2 | 208.21 | 208.22 | 208.8 | 208.81 | 208.82 | 208.9 | 208.91 | 208.92 | 196 |
| 196.1 | 196.2 | 196.3 | 196.5 | 196.6 | 196.8 | 196.9 | 197 | 197.1 | 197.2 | 197.3 |
| 197.4 | 197.5 | 197.6 | 197.7 | 197.8 | 198 | 198.1 | 198.2 | 198.3 | 198.4 | 198.5 |
| 198.6 | 198.7 | 198.8 | 198.81 | 198.82 | 198.89 | 199 | 199.1 | | | |

**Supplementary Table 2. Formula of the variability measures**

| Variability measure | Definition |
|---|---|
| Standard deviation | $\text{SD}=\sqrt{\frac{\sum \lvert x-\bar{x}\rvert^2}{N}}$ |

**Supplementary Table 3. The baseline, and clinical characteristics of patients in the study cohorts of four quartiles remnant cholesterol.**
* for p≤ 0.05, ** for p ≤ 0.01, *** for p ≤ 0.001, SD: standard deviation, ACEI: angiotensin-converting-enzyme inhibitors, ARB: angiotensin II receptor blockers, eGFR: estimated glomerular filtration rate.

| **Characteristics** | **All (N=75342) Mean(SD);N or Count(%)** | **Remnant cholesterol, mmol/L (Q1) (N=18779) Mean(SD);N or Count(%)** | **Remnant cholesterol, mmol/L (Q2) (N=18964) Mean(SD);N or Count(%)** | **Remnant cholesterol, mmol/L (Q3) (N=18838) Mean(SD);N or Count(%)** | **Remnant cholesterol, mmol/L (Q4) (N=18761) Mean(SD);N or Count(%)** | **P-value** |
|---|---|---|---|---|---|---|
| Male gender | 29905(39.69%) | 7697(40.98%) | 7519(39.64%) | 7245(38.45%) | 7444(39.67%) | <0.001** |
| Female gender | 45437(60.30%) | 11082(59.01%) | 11445(60.35%) | 11593(61.54%) | 11317(60.32%) | <0.001** |
| Baseline age, year | 61.3(13.1);n=75342 | 60.3(13.8);n=18779 | 61.6(13.1);n=18964 | 61.6(12.7);n=18838 | 61.6(12.8);n=18761 | <0.001** |
| **Remnant cholesterol** | | | | | | |
| Remnant cholesterol, mmol/L | 0.8(0.5);n=75342 | 0.3(0.1);n=18779 | 0.5(0.1);n=18964 | 0.78(0.09);n=18838 | 1.5(0.7);n=18761 | <0.001** |
| Time weighted remnant cholesterol, mmol/L | 0.9(0.8);n=75342 | 0.8(0.8);n=18779 | 0.8(0.7);n=18964 | 0.88(0.73);n=18838 | 1.1(0.9);n=18761 | <0.001** |
| **Comorbidities** | | | | | | |
| Charlson's standard comorbidity index | 1.7(1.3);n=75342 | 1.7(1.3);n=18779 | 1.8(1.3);n=18964 | 1.8(1.2);n=18838 | 1.8(1.2);n=18761 | <0.001** |
| Dyslipidaemia | 1288(1.70%) | 188(1.00%) | 280(1.47%) | 348(1.84%) | 472(2.51%) | <0.001** |
| Diabetes mellitus | 7889(10.47%) | 1395(7.42%) | 1638(8.63%) | 2064(10.95%) | 2792(14.88%) | <0.001** |

| | | | | | | |
|---|---|---|---|---|---|---|
| Hypertension | 39026(51.79%) | 8800(46.86%) | 9815(51.75%) | 10104(53.63%) | 10307(54.93%) | <0.001** |
| Acute myocardial infarction | 173(0.22%) | 30(0.15%) | 48(0.25%) | 48(0.25%) | 47(0.25%) | 0.149 |
| Ischemic heart disease | 1516(2.01%) | 322(1.71%) | 362(1.90%) | 395(2.09%) | 437(2.32%) | <0.001** |
| Heart failure | 2932(3.89%) | 557(2.96%) | 652(3.43%) | 784(4.16%) | 939(5.00%) | <0.001** |
| Atrial fibrillation/Atrial flutter | 446(0.59%) | 116(0.61%) | 120(0.63%) | 106(0.56%) | 104(0.55%) | 0.688 |
| Cerebrovascular accident, stroke | 545(0.72%) | 102(0.54%) | 144(0.75%) | 146(0.77%) | 153(0.81%) | 0.008* |
| Mental health diseases | 668(0.88%) | 144(0.76%) | 168(0.88%) | 160(0.84%) | 196(1.04%) | 0.034* |
| Chronic obstructive pulmonary disease | 189(0.25%) | 66(0.35%) | 57(0.30%) | 38(0.20%) | 28(0.14%) | <0.001** |
| Gastrointestinal bleeding | 461(0.61%) | 110(0.58%) | 111(0.58%) | 119(0.63%) | 121(0.64%) | 0.828 |
| Other bleeding | 982(1.30%) | 282(1.50%) | 251(1.32%) | 248(1.31%) | 201(1.07%) | 0.003* |
| Hip fractures | 419(0.55%) | 105(0.55%) | 100(0.52%) | 122(0.64%) | 92(0.49%) | 0.203 |
| History of falls | 1047(1.38%) | 249(1.32%) | 263(1.38%) | 282(1.49%) | 253(1.34%) | 0.499 |
| Accident fall | 1044(1.38%) | 249(1.32%) | 262(1.38%) | 281(1.49%) | 252(1.34%) | 0.515 |
| Liver diseases | 269(0.35%) | 87(0.46%) | 68(0.35%) | 56(0.29%) | 58(0.30%) | 0.028* |
| Renal diseases | 105(0.13%) | 26(0.13%) | 18(0.09%) | 31(0.16%) | 30(0.15%) | 0.249 |
| Cancer | 683(0.90%) | 177(0.94%) | 180(0.94%) | 176(0.93%) | 150(0.79%) | 0.361 |
| **Medications** | | | | | | |
| ACEI/ARB | 8890(11.79%) | 1755(9.34%) | 2100(11.07%) | 2282(12.11%) | 2753(14.67%) | <0.001** |

| | | | | | | |
|---|---|---|---|---|---|---|
| Anticoagulants | 37246(49.43%) | 8143(43.36%) | 9028(47.60%) | 9662(51.28%) | 10413(55.50%) | <0.001** |
| Antiplatelets | 7921(10.51%) | 1662(8.85%) | 1966(10.36%) | 2051(10.88%) | 2242(11.95%) | <0.001** |
| Lipid-lowering drugs | 7021(9.31%) | 1200(6.39%) | 1624(8.56%) | 1840(9.76%) | 2357(12.56%) | <0.001** |
| Statins and fibrates | 8460(11.22%) | 1353(7.20%) | 1825(9.62%) | 2207(11.71%) | 3075(16.39%) | <0.001** |
| Nitrates | 6148(8.16%) | 1265(6.73%) | 1548(8.16%) | 1544(8.19%) | 1791(9.54%) | <0.001** |
| Diuretics for heart failure | 1738(2.30%) | 396(2.10%) | 418(2.20%) | 425(2.25%) | 499(2.65%) | 0.002* |
| Diuretics for hypertension | 13805(18.32%) | 2610(13.89%) | 3273(17.25%) | 3655(19.40%) | 4267(22.74%) | <0.001** |
| Calcium channel blockers | 15126(20.07%) | 3090(16.45%) | 3581(18.88%) | 4086(21.69%) | 4369(23.28%) | <0.001** |
| Beta blockers | 15006(19.91%) | 2792(14.86%) | 3503(18.47%) | 4042(21.45%) | 4669(24.88%) | <0.001** |
| Non-steroidal anti-inflammatory drugs | 7594(10.07%) | 1604(8.54%) | 1890(9.96%) | 1964(10.42%) | 2136(11.38%) | <0.001** |
| Anti-diabetic drugs | 15121(20.06%) | 2860(15.22%) | 3483(18.36%) | 3996(21.21%) | 4782(25.48%) | <0.001** |
| **Laboratory testing** | | | | | | |
| eGFR (MDRD), mL/min/1.73m2 | 73.7(19.0);n=42133 | 76.2(19.3);n=9633 | 74.1(18.8);n=9594 | 73.2(18.8);n=10493 | 71.9(18.8);n=12413 | <0.001** |
| Urea, mmol/L | 5.9(2.1);n=41876 | 5.8(2.2);n=9570 | 5.9(2.19);n=9544 | 5.88(2.07);n=10426 | 5.94(2.17);n=12336 | <0.001** |
| Creatinine, umol/L | 87.9(29.7);n=42133 | 85.7(29.3);n=9633 | 87.4(30.0);n=9594 | 88.0(26.7);n=10493 | 89.7(31.9);n=12413 | <0.001** |

| | | | | | | |
|---|---|---|---|---|---|---|
| Aspartate transaminase, U/L | 28.9(59.0);n=16331 | 26.8(60.8);n=3698 | 27.3(43.6);n=3684 | 29.5(58.3);n=4037 | 31.0(67.6);n=4912 | <0.001** |
| Alanine transaminase, U/L | 28.4(56.8);n=18912 | 27.8(90.0);n=4212 | 26.9(43.6);n=4157 | 28.7(46.5);n=4552 | 29.6(39.2);n=5991 | <0.001** |
| **Blood pressures** | | | | | | |
| Systolic blood pressure, mmHg | 139.3(21.1);n=73878 | 136.6(21.5);n=18436 | 139.6(21.1);n=18610 | 140.4(20.7);n=18459 | 140.6(20.7);n=18373 | <0.001** |
| SD of systolic blood pressure | 13.7(5.8);n=70357 | 13.2(5.8);n=17604 | 13.68(5.74);n=17764 | 13.8(5.7);n=17582 | 14.0(5.8);n=17407 | <0.001** |
| Diastolic blood pressure, mmHg | 76.3(11.4);n=73878 | 75.2(11.4);n=18436 | 76.3(11.4);n=18610 | 76.8(11.3);n=18459 | 77.1(11.5);n=18373 | <0.001** |
| **Diabetes profile** | | | | | | |
| HbA1C, % | 7.5(1.7);n=12208 | 7.3(1.7);n=2329 | 7.4(1.7);n=2602 | 7.52(1.72);n=3180 | 7.7(1.7);n=4097 | <0.001** |
| SD of HbA1C | 0.7(0.7);n=8688 | 0.68(0.65);n=1615 | 0.71(0.69);n=1844 | 0.74(0.73);n=2290 | 0.8(0.7);n=2939 | <0.001** |
| Time weighted mean HbA1C, % | 7.2(1.2);n=13251 | 7.2(1.26);n=2528 | 7.22(1.22);n=2938 | 7.22(1.32);n=3502 | 7.3(1.2);n=4283 | 0.077 |
| Fasting glucose, mmol/L | 6.9(3.0);n=26975 | 6.6(2.9);n=5737 | 6.7(2.8);n=6052 | 7.0(3.0);n=6816 | 7.2(3.2);n=8370 | <0.001** |
| SD of fasting glucose | 1.3(1.6);n=13653 | 1.2(1.5);n=2661 | 1.2(1.6);n=2905 | 1.25(1.51);n=3394 | 1.3(1.6);n=4693 | <0.001** |
| Time weighted mean fasting glucose, mmol/L | 6.7(2.4);n=21939 | 6.5(2.5);n=4294 | 6.6(2.5);n=4859 | 6.8(2.5);n=5690 | 7.0(2.3);n=7096 | <0.001** |
| **Lipid profile** | | | | | | |
| Triglyceride, mmol/L | 1.6(1.1);n=75342 | 0.8(0.4);n=18779 | 1.2(0.3);n=18964 | 1.7(0.4);n=18838 | 2.7(1.6);n=18761 | <0.001** |
| SD of triglyceride | 0.5(0.5);n=75022 | 0.3(0.3);n=18695 | 0.3(0.3);n=18885 | 0.46(0.4);n=18751 | 0.8(0.8);n=18691 | <0.001** |

| | | | | | | |
|---|---|---|---|---|---|---|
| Time weighted mean triglyceride, mmol/L | 1.5(0.5);n=75274 | 1.3(0.4);n=18755 | 1.4(0.4);n=18955 | 1.6(0.4);n=18823 | 1.8(0.6);n=18741 | <0.001** |
| Low-density lipoprotein, mmol/L | 3.2(0.9);n=75342 | 3.0(0.8);n=18779 | 3.2(0.9);n=18964 | 3.3(0.9);n=18838 | 3.1(1.0);n=18761 | <0.001** |
| SD of low-density lipoprotein | 0.6(0.3);n=73404 | 0.5(0.3);n=18348 | 0.58(0.31);n=18589 | 0.62(0.32);n=18409 | 0.64(0.32);n=18058 | <0.001** |
| Time weighted mean low-density lipoprotein, mmol/L | 2.7(0.5);n=73445 | 2.68(0.48);n=18347 | 2.72(0.47);n=18621 | 2.73(0.48);n=18454 | 2.7(0.51);n=18023 | <0.001** |
| High-density lipoprotein, mmol/L | 1.3(0.4);n=75342 | 1.6(0.4);n=18779 | 1.4(0.3);n=18964 | 1.3(0.3);n=18838 | 1.2(0.3);n=18761 | <0.001** |
| SD of high-density lipoprotein | 0.2(0.1);n=71219 | 0.18(0.11);n=17824 | 0.17(0.09);n=18067 | 0.16(0.09);n=17823 | 0.15(0.09);n=17505 | <0.001** |
| Time weighted mean high-density lipoprotein, mmol/L | 1.3(0.2);n=71256 | 1.4(0.2);n=17820 | 1.4(0.2);n=18104 | 1.3(0.19);n=17869 | 1.3(0.2);n=17463 | <0.001** |
| Total cholesterol, mmol/L | 5.3(1.0);n=75342 | 4.9(0.9);n=18779 | 5.2(1.0);n=18964 | 5.4(1.0);n=18838 | 5.7(1.1);n=18761 | <0.001** |
| SD of total cholesterol | 0.7(0.4);n=75061 | 0.6(0.3);n=18716 | 0.6(0.3);n=18884 | 0.69(0.35);n=18756 | 0.8(0.4);n=18705 | <0.001** |
| Time weighted mean total cholesterol, mmol/L | 4.8(0.5);n=75342 | 4.7(0.6);n=18779 | 4.76(0.54);n=18964 | 4.8(0.53);n=18838 | 4.84(0.54);n=18761 | <0.001** |

**Supplementary Table 4. The outcome of patients in the study cohorts of four quartiles remnant cholesterol.**

IQR: interquartile range

| Characteristics | All (N=75342) Median (IQR);N or Count(%) | Remnant cholesterol, mmol/L (Q1) (N=18779) Median (IQR);N or Count(%) | Remnant cholesterol, mmol/L (Q2) (N=18964) Median (IQR);N or Count(%) | Remnant cholesterol, mmol/L (Q3) (N=18838) Median (IQR);N or Count(%) | Remnant cholesterol, mmol/L (Q4) (N=18761) Median (IQR);N or Count(%) | P-value |
|---|---|---|---|---|---|---|
| All-cause mortality | 23475(31.15%) | 5451(29.02%) | 5814(30.65%) | 5925(31.45%) | 6285(33.50%) | <0.001** |
| Cancer-related mortality | 4349(5.77%) | 994(5.29%) | 1104(5.82%) | 1118(5.93%) | 1133(6.03%) | <0.001** |
| Cardiovascular-related mortality | 4533(6.01%) | 1006(5.35%) | 1104(5.82%) | 1108(5.88%) | 1315(7.00%) | <0.001** |
| Ischaemic heart disease mortality | 1925(2.55%) | 384(2.04%) | 470(2.47%) | 483(2.56%) | 588(3.13%) | <0.001** |
| Respiratory-related mortality | 7424(9.85%) | 1913(10.18%) | 1911(10.07%) | 1805(9.58%) | 1795(9.56%) | <0.001** |
| Pneumonia-related mortality | 6388(8.47%) | 1642(8.74%) | 1655(8.72%) | 1559(8.27%) | 1532(8.16%) | <0.001** |
| Chronic lower respiratory related mortality | 250(0.33%) | 90(0.47%) | 69(0.36%) | 40(0.21%) | 51(0.27%) | <0.001** |
| Gastrointestinal-related mortality | 663(0.87%) | 144(0.76%) | 162(0.85%) | 195(1.03%) | 162(0.86%) | <0.001** |
| Infection-related mortality | 780(1.03%) | 168(0.89%) | 185(0.97%) | 203(1.07%) | 224(1.19%) | <0.001** |
| Genitourinary-related mortality | 1775(2.35%) | 290(1.54%) | 387(2.04%) | 486(2.57%) | 612(3.26%) | <0.001** |
| Cerebrovascular-related mortality | 1265(1.67%) | 313(1.66%) | 310(1.63%) | 295(1.56%) | 347(1.84%) | <0.001** |
| Kidney-related mortality | 1351(1.79%) | 215(1.14%) | 287(1.51%) | 368(1.95%) | 481(2.56%) | <0.001** |
| Septicaemia-related mortality | 676(0.89%) | 130(0.69%) | 160(0.84%) | 179(0.95%) | 207(1.10%) | <0.001** |
| Diabetes mellitus related mortality | 64(0.08%) | 9(0.04%) | 12(0.06%) | 17(0.09%) | 26(0.13%) | 0.002* |
| Haematological and endocrinological-related mortality | 223(0.29%) | 50(0.26%) | 43(0.22%) | 66(0.35%) | 64(0.34%) | <0.001** |
| Injury, poisoning and certain other consequences of external causes | 309(0.41%) | 83(0.44%) | 97(0.51%) | 66(0.35%) | 63(0.33%) | 0.022* |
| Unknown mortality | 2627(3.48%) | 592(3.15%) | 654(3.44%) | 679(3.60%) | 702(3.74%) | <0.001** |

**Supplementary Figure 1. Distribution and association analysis between remnant cholesterol and HDL-C, LDL-C, TC, and TG.**

HDL-C = high density lipoprotein cholesterol, LDL-C = low density lipoprotein cholesterol, RR = risk ratios, TC = total cholesterol, TG = triglyceride

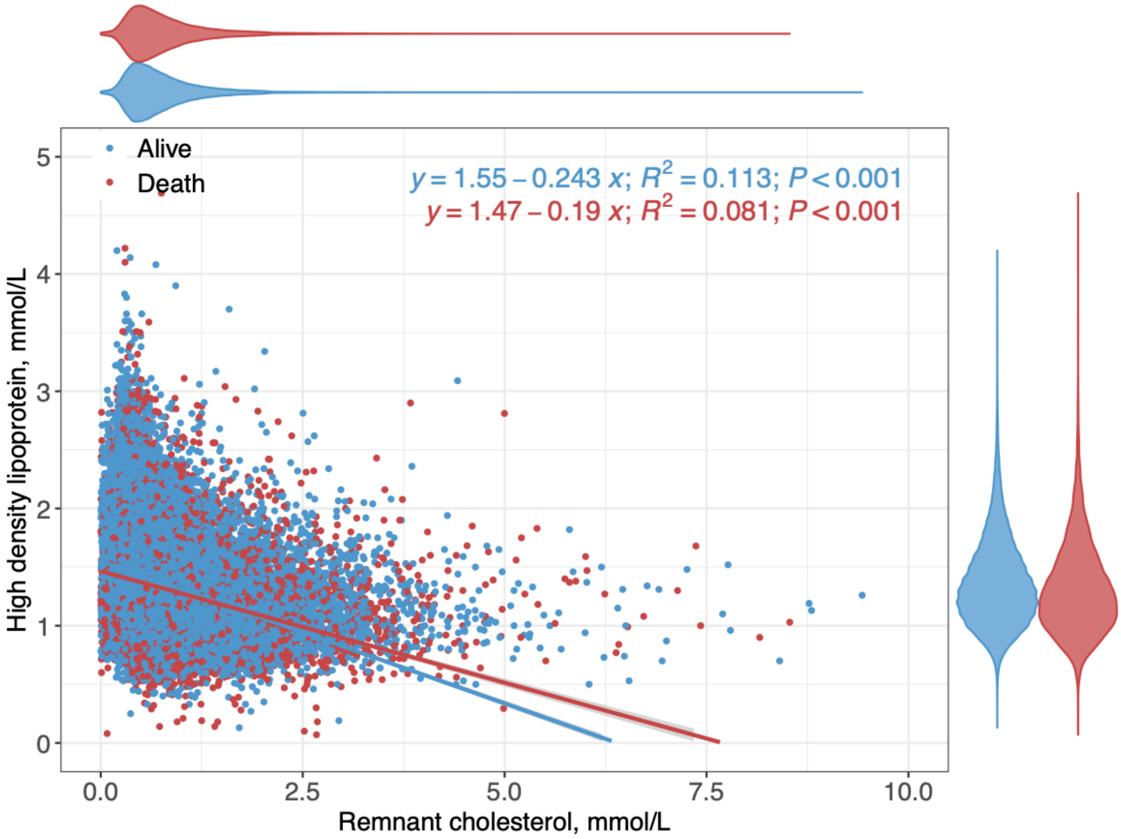

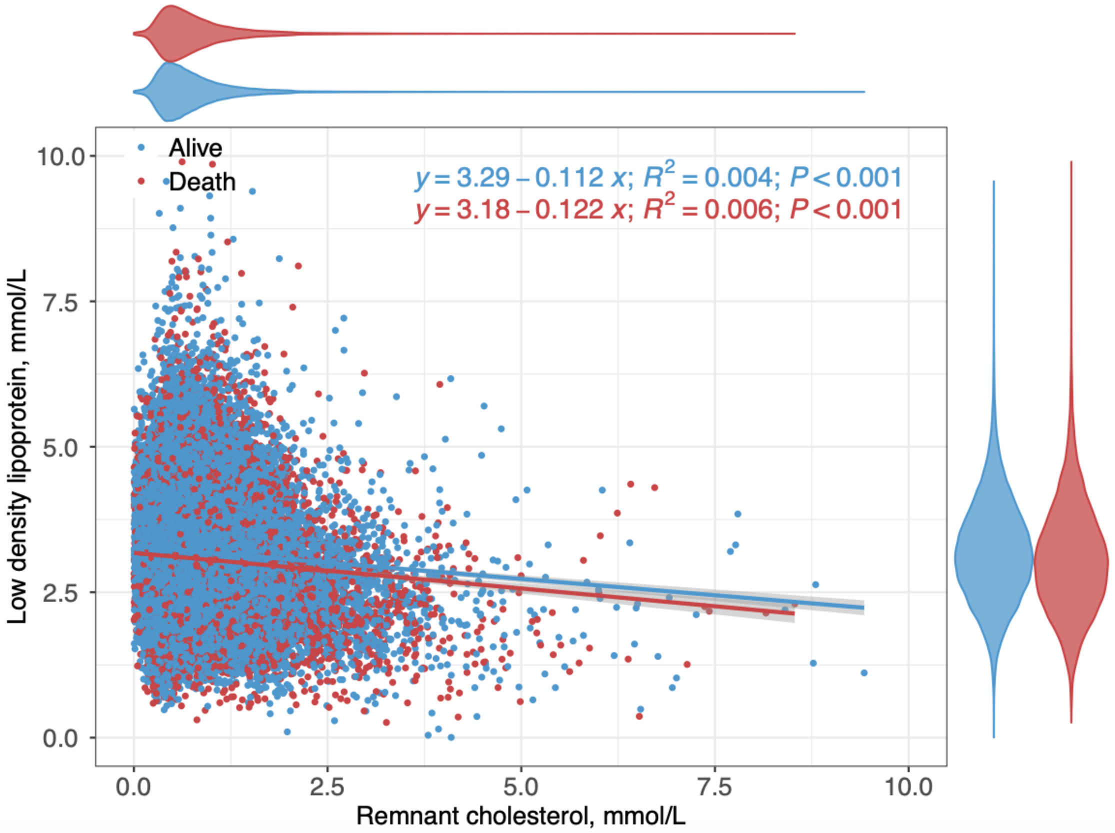

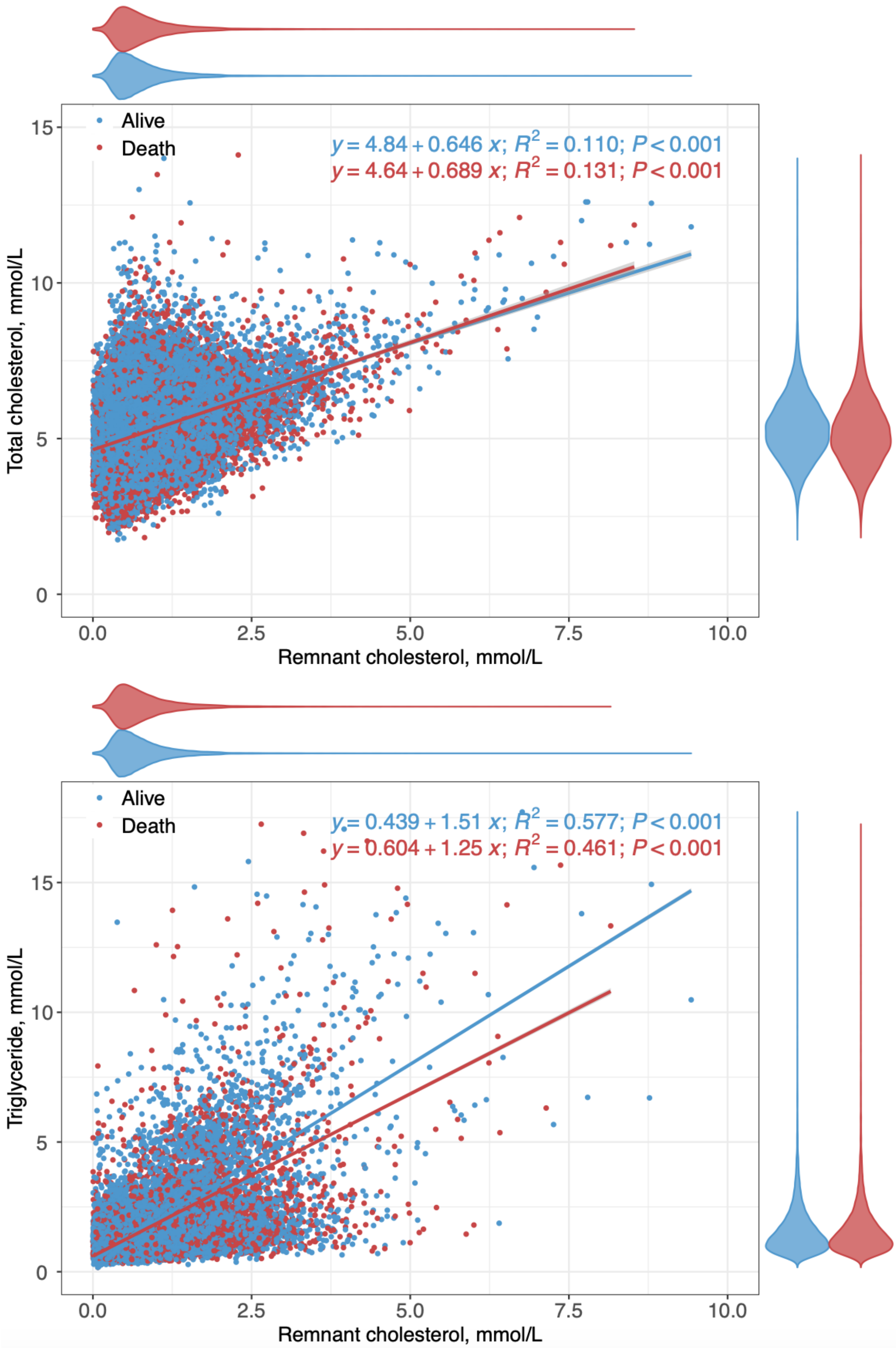

Alive
Death
y = 4.84 + 0.646 x; R2 = 0.110; P < 0.001
y = 4.64 + 0.689 x; R2 = 0.131; P < 0.001
Total cholesterol, mmol/L
Remnant cholesterol, mmol/L
Alive
Death
y = 0.439 + 1.51 x; R2 = 0.577; P < 0.001
y = 0.604 + 1.25 x; R2 = 0.461; P < 0.001
Triglyceride, mmol/L
Remnant cholesterol, mmol/L

**Supplementary Figure 2. Cumulative incidence curves stratified by quartiles of remnant cholesterol to predict mortality outcome.**

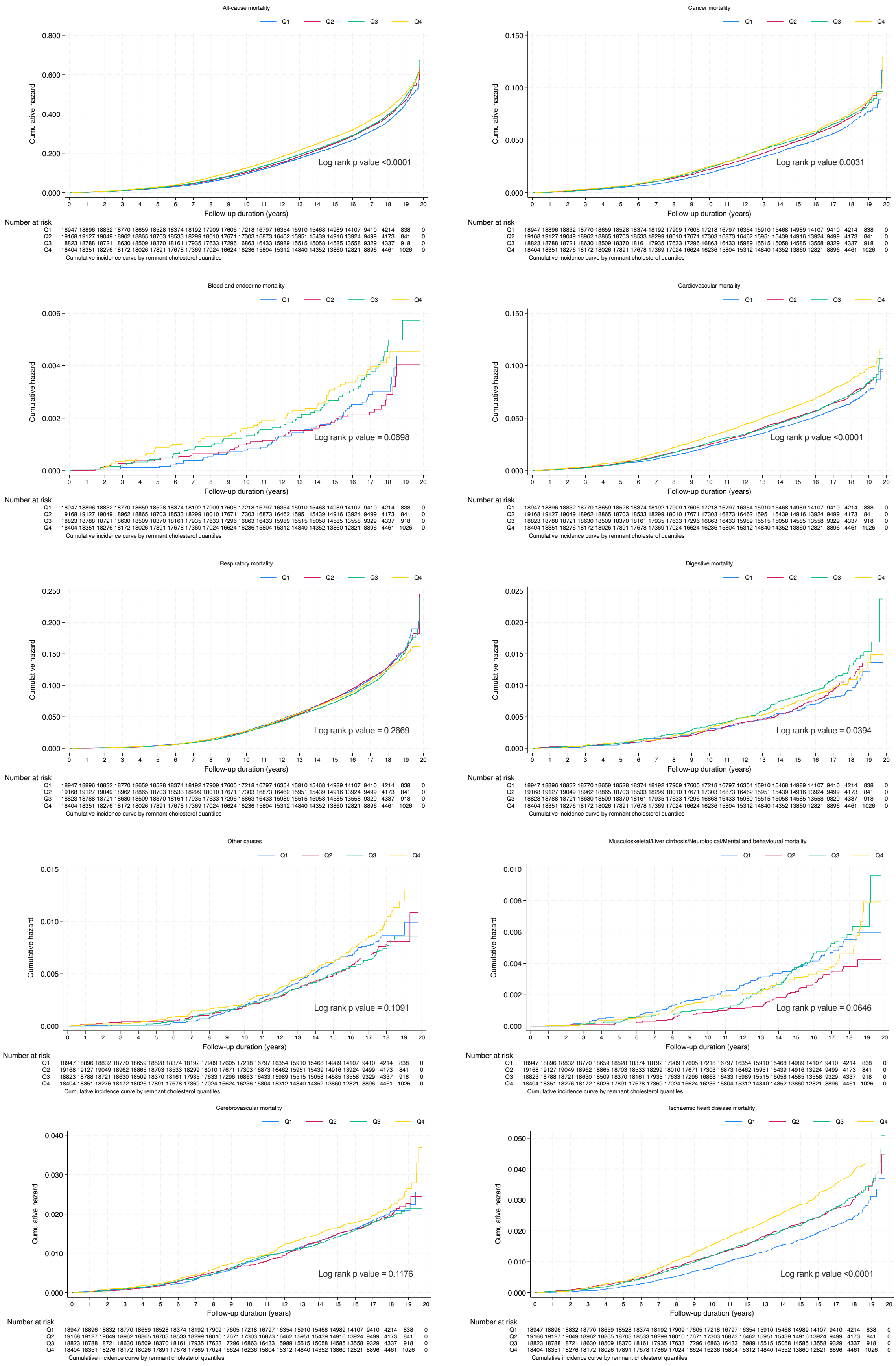

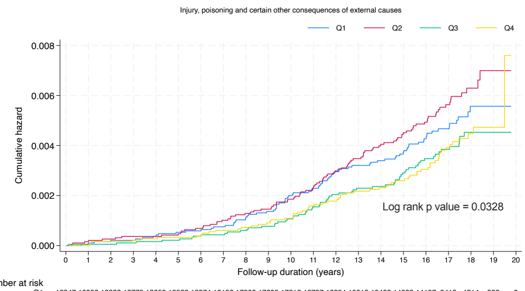

Number at risk

| | | | | | | | | | | | | | | | | | | | | | |
|---|---|---|---|---|---|---|---|---|---|---|---|---|---|---|---|---|---|---|---|---|---|
| Q1 | 18947 | 18896 | 18832 | 18770 | 18659 | 18528 | 18374 | 18192 | 17909 | 17605 | 17218 | 16797 | 16354 | 15910 | 15468 | 14989 | 14107 | 9410 | 4214 | 838 | 0 |
| Q2 | 19168 | 19127 | 19049 | 18962 | 18865 | 18703 | 18533 | 18299 | 18010 | 17671 | 17303 | 16873 | 16462 | 15951 | 15439 | 14916 | 13924 | 9499 | 4173 | 841 | 0 |
| Q3 | 18823 | 18788 | 18721 | 18630 | 18509 | 18370 | 18161 | 17935 | 17633 | 17296 | 16863 | 16433 | 15989 | 15515 | 15058 | 14585 | 13558 | 9329 | 4337 | 918 | 0 |
| Q4 | 18404 | 18351 | 18276 | 18172 | 18026 | 17891 | 17678 | 17369 | 17024 | 16624 | 16236 | 15804 | 15312 | 14840 | 14352 | 13860 | 12821 | 8896 | 4461 | 1026 | 0 |

Cumulative incidence curve by remnant cholesterol quantiles

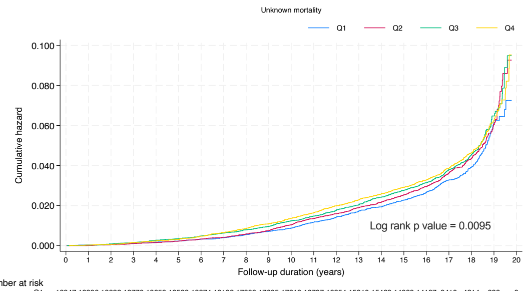

Number at risk

| | | | | | | | | | | | | | | | | | | | | | |
|---|---|---|---|---|---|---|---|---|---|---|---|---|---|---|---|---|---|---|---|---|---|
| Q1 | 18947 | 18896 | 18832 | 18770 | 18659 | 18528 | 18374 | 18192 | 17909 | 17605 | 17218 | 16797 | 16354 | 15910 | 15468 | 14989 | 14107 | 9410 | 4214 | 838 | 0 |
| Q2 | 19168 | 19127 | 19049 | 18962 | 18865 | 18703 | 18533 | 18299 | 18010 | 17671 | 17303 | 16873 | 16462 | 15951 | 15439 | 14916 | 13924 | 9499 | 4173 | 841 | 0 |
| Q3 | 18823 | 18788 | 18721 | 18630 | 18509 | 18370 | 18161 | 17935 | 17633 | 17296 | 16863 | 16433 | 15989 | 15515 | 15058 | 14585 | 13558 | 9329 | 4337 | 918 | 0 |
| Q4 | 18404 | 18351 | 18276 | 18172 | 18026 | 17891 | 17678 | 17369 | 17024 | 16624 | 16236 | 15804 | 15312 | 14840 | 14352 | 13860 | 12821 | 8896 | 4461 | 1026 | 0 |

Cumulative incidence curve by remnant cholesterol quantiles

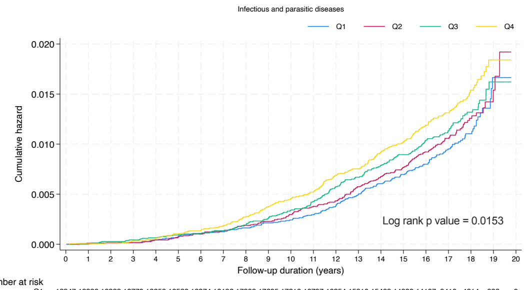

Number at risk

| | | | | | | | | | | | | | | | | | | | | | |
|---|---|---|---|---|---|---|---|---|---|---|---|---|---|---|---|---|---|---|---|---|---|
| Q1 | 18947 | 18896 | 18832 | 18770 | 18659 | 18528 | 18374 | 18192 | 17909 | 17605 | 17218 | 16797 | 16354 | 15910 | 15468 | 14989 | 14107 | 9410 | 4214 | 838 | 0 |
| Q2 | 19168 | 19127 | 19049 | 18962 | 18865 | 18703 | 18533 | 18299 | 18010 | 17671 | 17303 | 16873 | 16462 | 15951 | 15439 | 14916 | 13924 | 9499 | 4173 | 841 | 0 |
| Q3 | 18823 | 18788 | 18721 | 18630 | 18509 | 18370 | 18161 | 17935 | 17633 | 17296 | 16863 | 16433 | 15989 | 15515 | 15058 | 14585 | 13558 | 9329 | 4337 | 918 | 0 |
| Q4 | 18404 | 18351 | 18276 | 18172 | 18026 | 17891 | 17678 | 17369 | 17024 | 16624 | 16236 | 15804 | 15312 | 14840 | 14352 | 13860 | 12821 | 8896 | 4461 | 1026 | 0 |

Cumulative incidence curve by remnant cholesterol quantiles

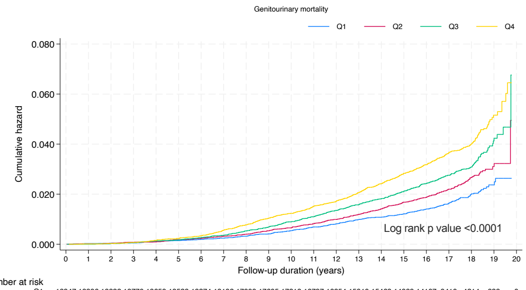

Number at risk

| | | | | | | | | | | | | | | | | | | | | | |
|---|---|---|---|---|---|---|---|---|---|---|---|---|---|---|---|---|---|---|---|---|---|
| Q1 | 18947 | 18896 | 18832 | 18770 | 18659 | 18528 | 18374 | 18192 | 17909 | 17605 | 17218 | 16797 | 16354 | 15910 | 15468 | 14989 | 14107 | 9410 | 4214 | 838 | 0 |
| Q2 | 19168 | 19127 | 19049 | 18962 | 18865 | 18703 | 18533 | 18299 | 18010 | 17671 | 17303 | 16873 | 16462 | 15951 | 15439 | 14916 | 13924 | 9499 | 4173 | 841 | 0 |
| Q3 | 18823 | 18788 | 18721 | 18630 | 18509 | 18370 | 18161 | 17935 | 17633 | 17296 | 16863 | 16433 | 15989 | 15515 | 15058 | 14585 | 13558 | 9329 | 4337 | 918 | 0 |
| Q4 | 18404 | 18351 | 18276 | 18172 | 18026 | 17891 | 17678 | 17369 | 17024 | 16624 | 16236 | 15804 | 15312 | 14840 | 14352 | 13860 | 12821 | 8896 | 4461 | 1026 | 0 |

Cumulative incidence curve by remnant cholesterol quantiles

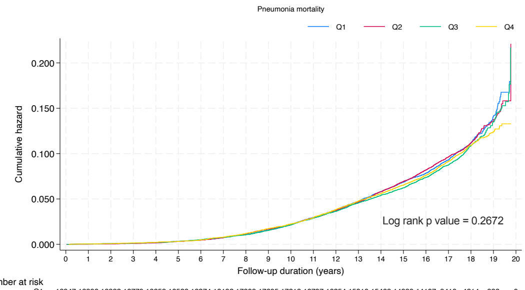

Number at risk

| | | | | | | | | | | | | | | | | | | | | | |
|---|---|---|---|---|---|---|---|---|---|---|---|---|---|---|---|---|---|---|---|---|---|
| Q1 | 18947 | 18896 | 18832 | 18770 | 18659 | 18528 | 18374 | 18192 | 17909 | 17605 | 17218 | 16797 | 16354 | 15910 | 15468 | 14989 | 14107 | 9410 | 4214 | 838 | 0 |
| Q2 | 19168 | 19127 | 19049 | 18962 | 18865 | 18703 | 18533 | 18299 | 18010 | 17671 | 17303 | 16873 | 16462 | 15951 | 15439 | 14916 | 13924 | 9499 | 4173 | 841 | 0 |
| Q3 | 18823 | 18788 | 18721 | 18630 | 18509 | 18370 | 18161 | 17935 | 17633 | 17296 | 16863 | 16433 | 15989 | 15515 | 15058 | 14585 | 13558 | 9329 | 4337 | 918 | 0 |
| Q4 | 18404 | 18351 | 18276 | 18172 | 18026 | 17891 | 17678 | 17369 | 17024 | 16624 | 16236 | 15804 | 15312 | 14840 | 14352 | 13860 | 12821 | 8896 | 4461 | 1026 | 0 |

Cumulative incidence curve by remnant cholesterol quantiles

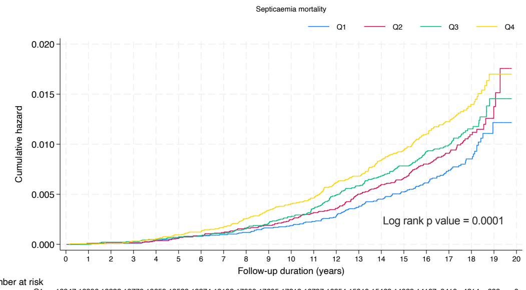

Number at risk

| | | | | | | | | | | | | | | | | | | | | | |
|---|---|---|---|---|---|---|---|---|---|---|---|---|---|---|---|---|---|---|---|---|---|
| Q1 | 18947 | 18896 | 18832 | 18770 | 18659 | 18528 | 18374 | 18192 | 17909 | 17605 | 17218 | 16797 | 16354 | 15910 | 15468 | 14989 | 14107 | 9410 | 4214 | 838 | 0 |
| Q2 | 19168 | 19127 | 19049 | 18962 | 18865 | 18703 | 18533 | 18299 | 18010 | 17671 | 17303 | 16873 | 16462 | 15951 | 15439 | 14916 | 13924 | 9499 | 4173 | 841 | 0 |
| Q3 | 18823 | 18788 | 18721 | 18630 | 18509 | 18370 | 18161 | 17935 | 17633 | 17296 | 16863 | 16433 | 15989 | 15515 | 15058 | 14585 | 13558 | 9329 | 4337 | 918 | 0 |
| Q4 | 18404 | 18351 | 18276 | 18172 | 18026 | 17891 | 17678 | 17369 | 17024 | 16624 | 16236 | 15804 | 15312 | 14840 | 14352 | 13860 | 12821 | 8896 | 4461 | 1026 | 0 |

Cumulative incidence curve by remnant cholesterol quantiles

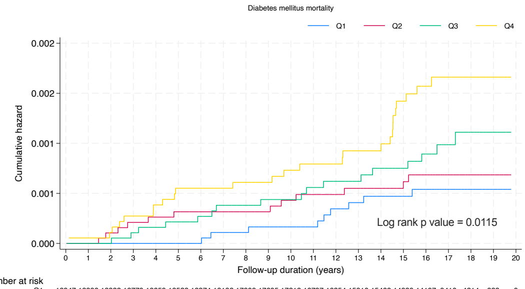

Number at risk

| | | | | | | | | | | | | | | | | | | | | | |
|---|---|---|---|---|---|---|---|---|---|---|---|---|---|---|---|---|---|---|---|---|---|
| Q1 | 18947 | 18896 | 18832 | 18770 | 18659 | 18528 | 18374 | 18192 | 17909 | 17605 | 17218 | 16797 | 16354 | 15910 | 15468 | 14989 | 14107 | 9410 | 4214 | 838 | 0 |
| Q2 | 19168 | 19127 | 19049 | 18962 | 18865 | 18703 | 18533 | 18299 | 18010 | 17671 | 17303 | 16873 | 16462 | 15951 | 15439 | 14916 | 13924 | 9499 | 4173 | 841 | 0 |
| Q3 | 18823 | 18788 | 18721 | 18630 | 18509 | 18370 | 18161 | 17935 | 17633 | 17296 | 16863 | 16433 | 15989 | 15515 | 15058 | 14585 | 13558 | 9329 | 4337 | 918 | 0 |
| Q4 | 18404 | 18351 | 18276 | 18172 | 18026 | 17891 | 17678 | 17369 | 17024 | 16624 | 16236 | 15804 | 15312 | 14840 | 14352 | 13860 | 12821 | 8896 | 4461 | 1026 | 0 |

Cumulative incidence curve by remnant cholesterol quantiles

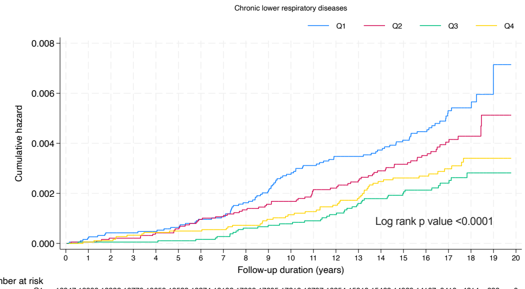

Number at risk

| | | | | | | | | | | | | | | | | | | | | | |
|---|---|---|---|---|---|---|---|---|---|---|---|---|---|---|---|---|---|---|---|---|---|
| Q1 | 18947 | 18896 | 18832 | 18770 | 18659 | 18528 | 18374 | 18192 | 17909 | 17605 | 17218 | 16797 | 16354 | 15910 | 15468 | 14989 | 14107 | 9410 | 4214 | 838 | 0 |
| Q2 | 19168 | 19127 | 19049 | 18962 | 18865 | 18703 | 18533 | 18299 | 18010 | 17671 | 17303 | 16873 | 16462 | 15951 | 15439 | 14916 | 13924 | 9499 | 4173 | 841 | 0 |
| Q3 | 18823 | 18788 | 18721 | 18630 | 18509 | 18370 | 18161 | 17935 | 17633 | 17296 | 16863 | 16433 | 15989 | 15515 | 15058 | 14585 | 13558 | 9329 | 4337 | 918 | 0 |
| Q4 | 18404 | 18351 | 18276 | 18172 | 18026 | 17891 | 17678 | 17369 | 17024 | 16624 | 16236 | 15804 | 15312 | 14840 | 14352 | 13860 | 12821 | 8896 | 4461 | 1026 | 0 |

Cumulative incidence curve by remnant cholesterol quantiles

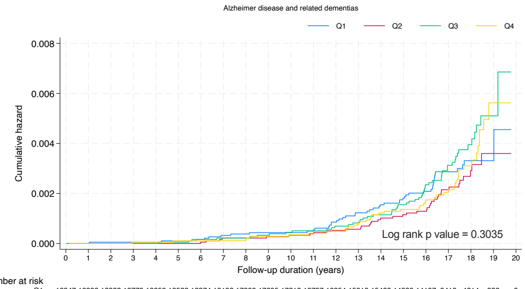

Number at risk

| | | | | | | | | | | | | | | | | | | | | | |
|---|---|---|---|---|---|---|---|---|---|---|---|---|---|---|---|---|---|---|---|---|---|
| Q1 | 18947 | 18896 | 18832 | 18770 | 18659 | 18528 | 18374 | 18192 | 17909 | 17605 | 17218 | 16797 | 16354 | 15910 | 15468 | 14989 | 14107 | 9410 | 4214 | 838 | 0 |
| Q2 | 19168 | 19127 | 19049 | 18962 | 18865 | 18703 | 18533 | 18299 | 18010 | 17671 | 17303 | 16873 | 16462 | 15951 | 15439 | 14916 | 13924 | 9499 | 4173 | 841 | 0 |
| Q3 | 18823 | 18788 | 18721 | 18630 | 18509 | 18370 | 18161 | 17935 | 17633 | 17296 | 16863 | 16433 | 15989 | 15515 | 15058 | 14585 | 13558 | 9329 | 4337 | 918 | 0 |
| Q4 | 18404 | 18351 | 18276 | 18172 | 18026 | 17891 | 17678 | 17369 | 17024 | 16624 | 16236 | 15804 | 15312 | 14840 | 14352 | 13860 | 12821 | 8896 | 4461 | 1026 | 0 |

Cumulative incidence curve by remnant cholesterol quantiles

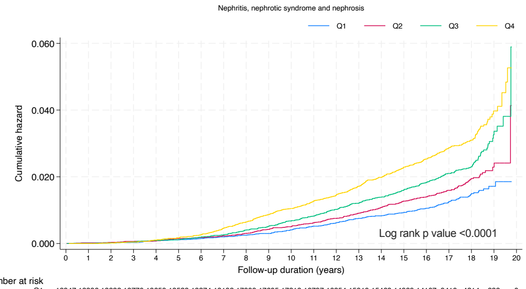

Number at risk

| | | | | | | | | | | | | | | | | | | | | | |
|---|---|---|---|---|---|---|---|---|---|---|---|---|---|---|---|---|---|---|---|---|---|
| Q1 | 18947 | 18896 | 18832 | 18770 | 18659 | 18528 | 18374 | 18192 | 17909 | 17605 | 17218 | 16797 | 16354 | 15910 | 15468 | 14989 | 14107 | 9410 | 4214 | 838 | 0 |
| Q2 | 19168 | 19127 | 19049 | 18962 | 18865 | 18703 | 18533 | 18299 | 18010 | 17671 | 17303 | 16873 | 16462 | 15951 | 15439 | 14916 | 13924 | 9499 | 4173 | 841 | 0 |
| Q3 | 18823 | 18788 | 18721 | 18630 | 18509 | 18370 | 18161 | 17935 | 17633 | 17296 | 16863 | 16433 | 15989 | 15515 | 15058 | 14585 | 13558 | 9329 | 4337 | 918 | 0 |
| Q4 | 18404 | 18351 | 18276 | 18172 | 18026 | 17891 | 17678 | 17369 | 17024 | 16624 | 16236 | 15804 | 15312 | 14840 | 14352 | 13860 | 12821 | 8896 | 4461 | 1026 | 0 |

Cumulative incidence curve by remnant cholesterol quantiles

**Supplementary Figure 3. Restricted cubic spline for remnant cholesterol predicting cause-related mortality on continuous scales.**

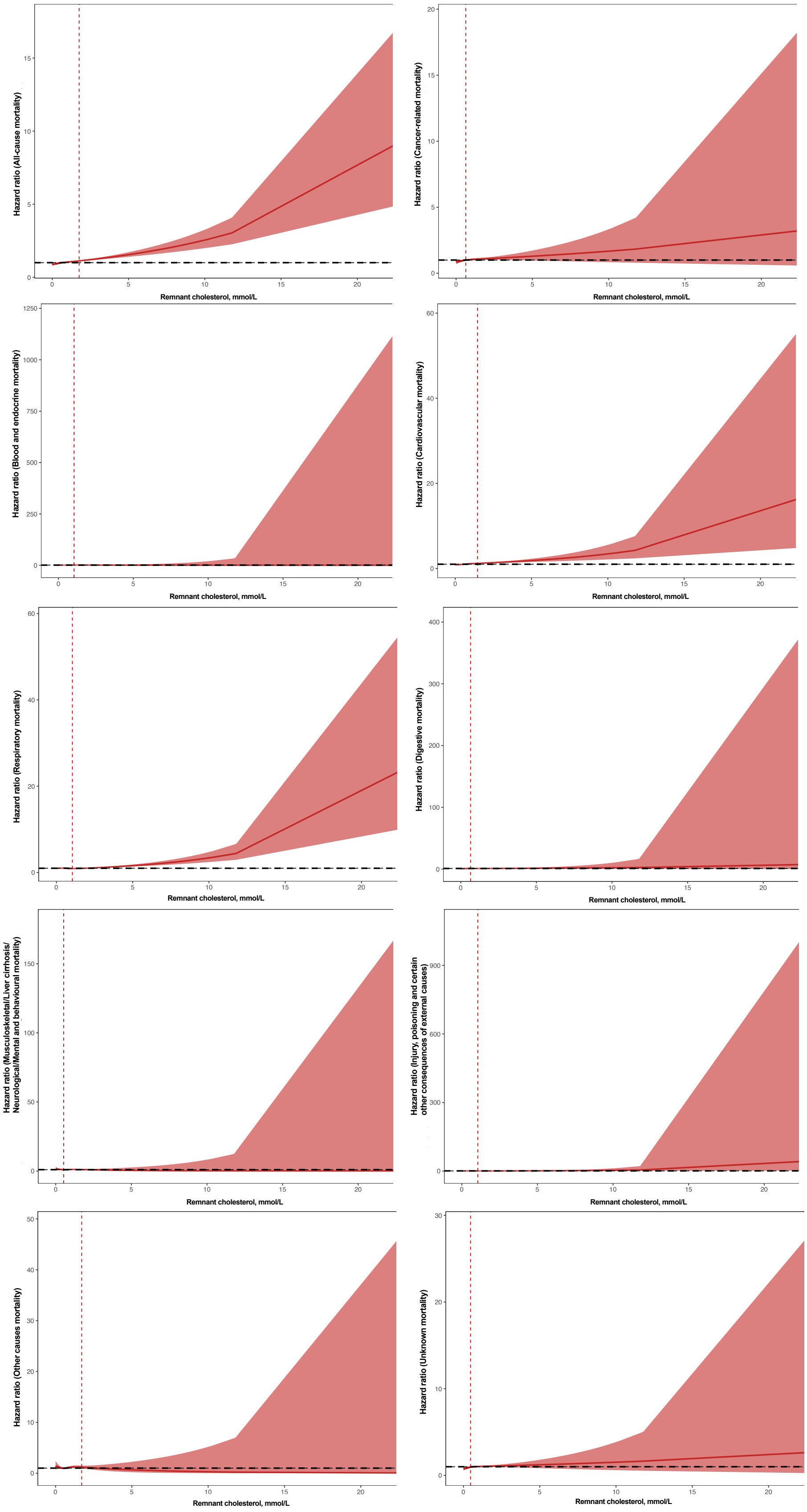

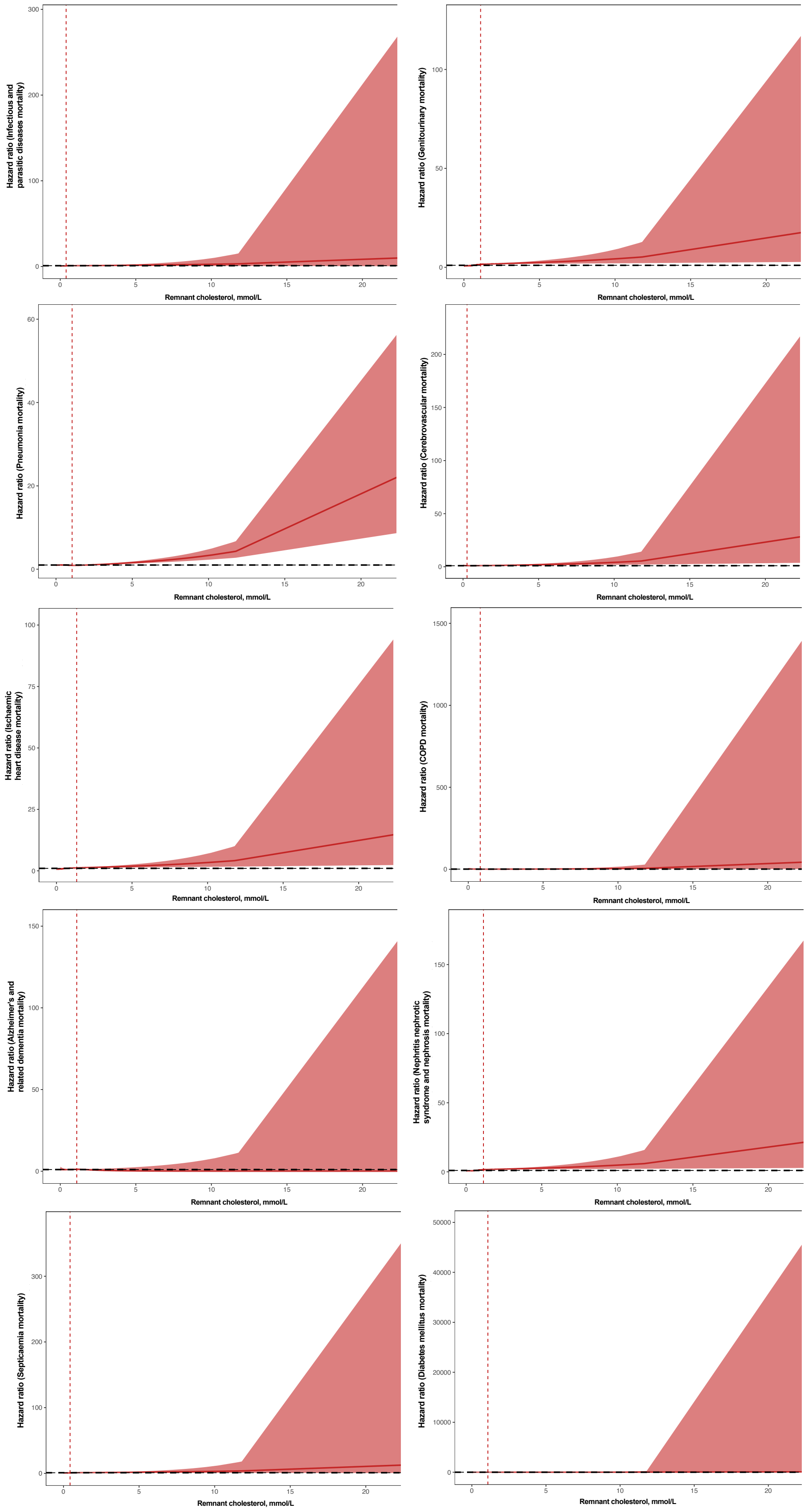
Hazard ratio (Infectious and parasitic diseases mortality)
Remnant cholesterol, mmol/L
Hazard ratio (Genitourinary mortality)
Remnant cholesterol, mmol/L
Hazard ratio (Pneumonia mortality)
Remnant cholesterol, mmol/L
Hazard ratio (Cerebrovascular mortality)
Remnant cholesterol, mmol/L
Hazard ratio (Ischaemic heart disease mortality)
Remnant cholesterol, mmol/L
Hazard ratio (COPD mortality)
Remnant cholesterol, mmol/L
Hazard ratio (Alzheimer's and related dementia mortality)
Remnant cholesterol, mmol/L
Hazard ratio (Nephritis nephrotic syndrome and nephrosis mortality)
Remnant cholesterol, mmol/L
Hazard ratio (Septicaemia mortality)
Remnant cholesterol, mmol/L
Hazard ratio (Diabetes mellitus mortality)
Remnant cholesterol, mmol/L

**Supplementary Figure 4. Restricted cubic spline comparing predicted cause-specific mortality between individuals with and without prior atherosclerotic cardiovascular disease.**

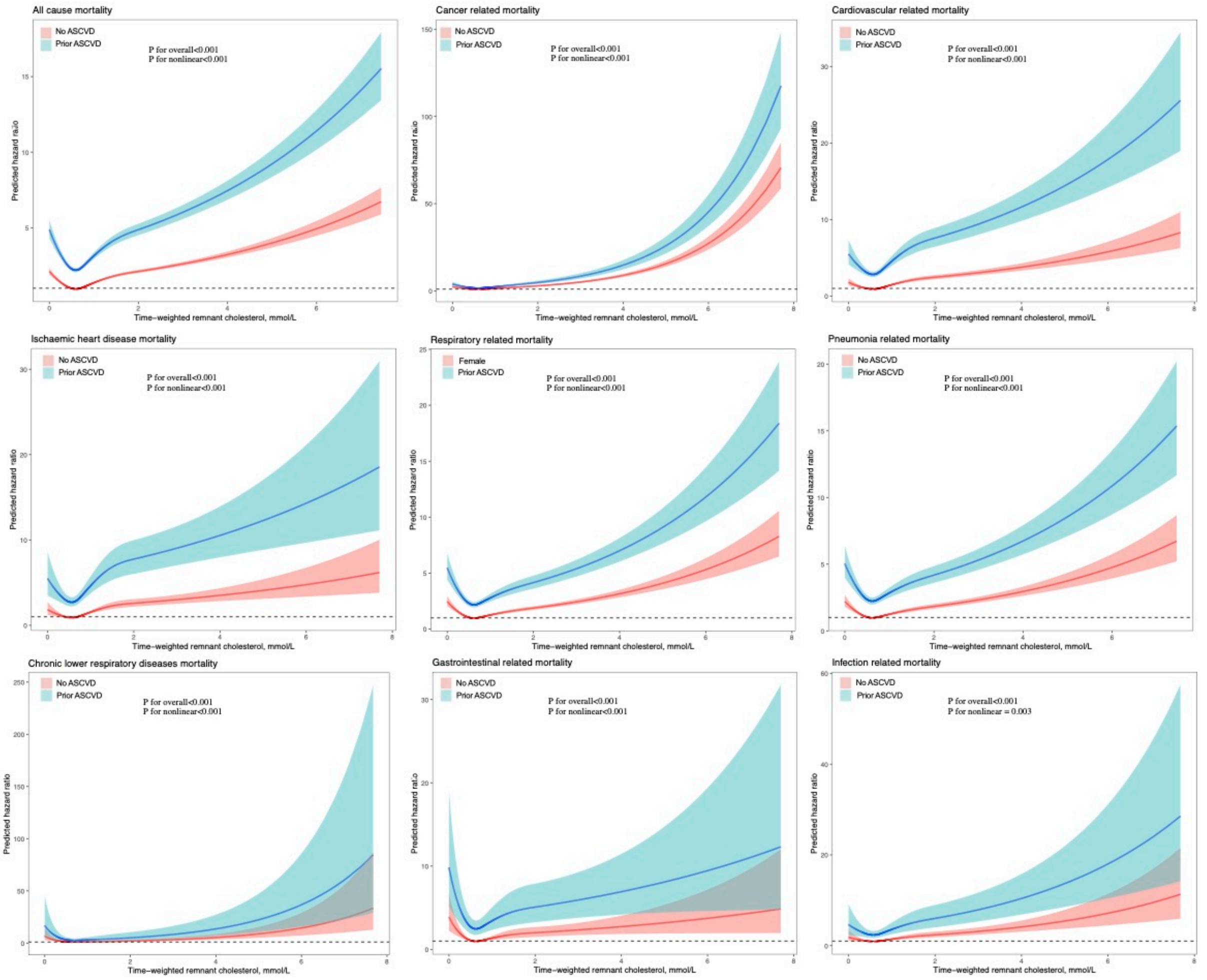

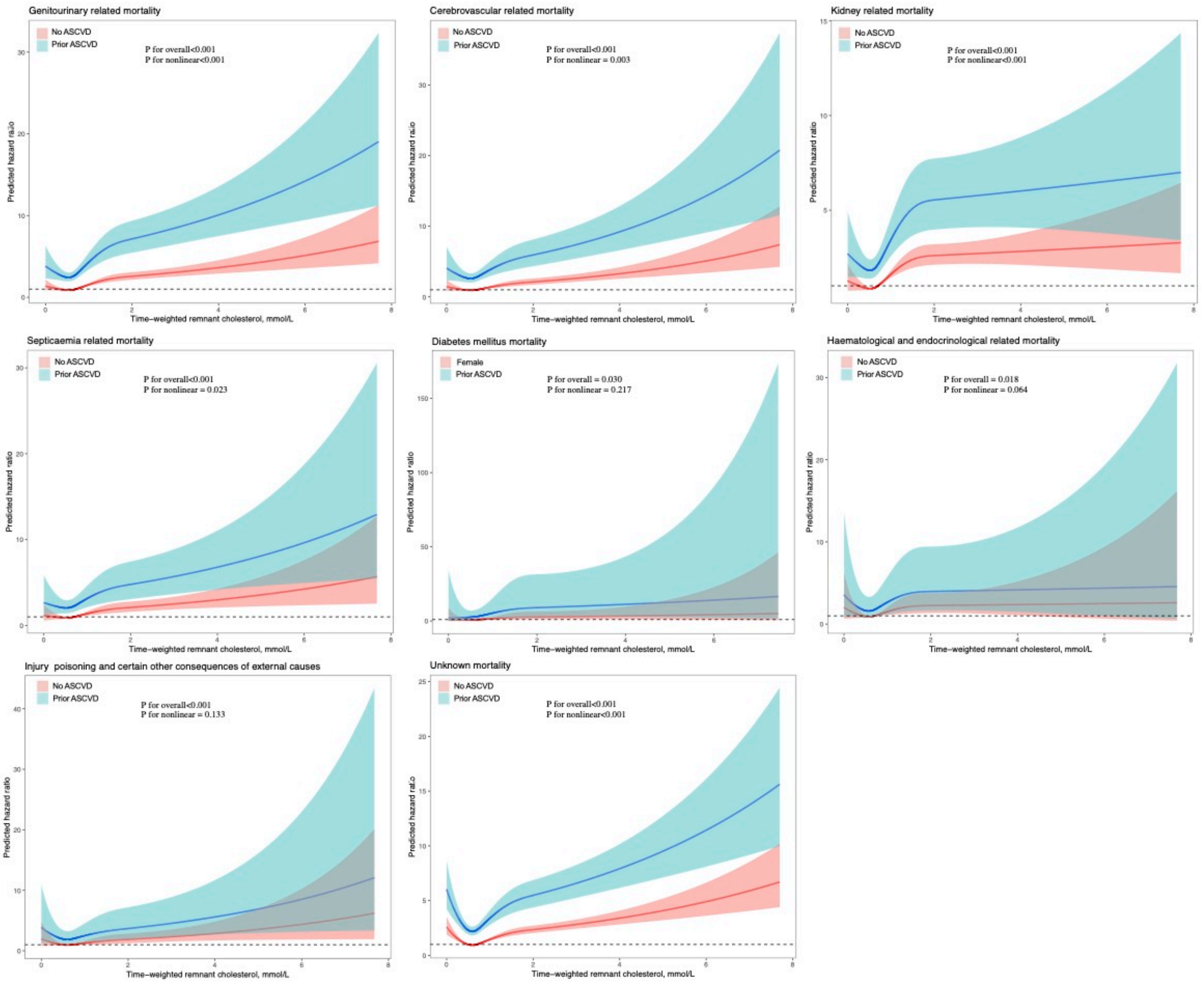
Genitourinary related mortality
No ASCVD
Prior ASCVD
P for overall<0.001
P for nonlinear<0.001
Cerebrovascular related mortality
No ASCVD
Prior ASCVD
P for overall<0.001
P for nonlinear = 0.003
Kidney related mortality
No ASCVD
Prior ASCVD
P for overall<0.001
P for nonlinear<0.001
Septicaemia related mortality
No ASCVD
Prior ASCVD
P for overall<0.001
P for nonlinear = 0.023
Diabetes mellitus mortality
Female
Prior ASCVD
P for overall = 0.030
P for nonlinear = 0.217
Haematological and endocrinological related mortality
No ASCVD
Prior ASCVD
P for overall = 0.018
P for nonlinear = 0.064
Injury poisoning and certain other consequences of external causes
No ASCVD
Prior ASCVD
P for overall<0.001
P for nonlinear = 0.133
Unknown mortality
No ASCVD
Prior ASCVD
P for overall<0.001
P for nonlinear<0.001
Predicted hazard ratio
Time−weighted remnant cholesterol, mmol/L
